# CenTauR: Towards a Universal Scale and Masks for Standardizing Tau Imaging Studies

**DOI:** 10.1101/2023.03.22.23287009

**Authors:** Victor L. Villemagne, Antoine Leuzy, Sandra Sanabria Bohorquez, Santiago Bullich, Hitoshi Shimada, Christopher C. Rowe, Pierrick Bourgeat, Brian Lopresti, Kun Huang, Natasha Krishnadas, Jurgen Fripp, Yuhei Takado, Alexandra Gogola, Davneet Minhas, Robby Weimer, Makoto Higuchi, Andrew Stephens, Oskar Hansson, Vincent Doré, the Alzheimer’s Disease Neuroimaging Initiative, the AIBL research group

## Abstract

**INTRODUCTION:** Recently, an increasing number of tau tracers have become available. There is a need to standardize quantitative tau measures across tracers, supporting a universal scale. We developed several cortical tau masks and applied them to generate a tau imaging universal scale.

**METHOD:** 1045 participants underwent tau scans with either ^18^F-Flortaucipir, ^18^F-MK6240, ^18^F-PI2620, ^18^F-PM-PBB3, ^18^F-GTP1 or ^18^F-RO948. The mask was generated from cognitively unimpaired Aβ-subjects and AD patients with Aβ+. Four additional regional cortical masks were defined within the constraints of the global mask. A universal scale, the CenTauR_z_, was constructed.

**RESULTS:** None of the regions known to display off-target signal were included in the masks. The CenTauR_z_ allows robustly discrimination between low and high levels of tau deposits.

**DISCUSSION:** We constructed several tau-specific cortical masks^*^ for the AD continuum and a universal standard scale designed to capture the location and degree of abnormality that can be applied across tracers and across centres.

**Research in Context:**

1. **Systematic review:** The authors reviewed the literature using traditional (e.g., PubMed) sources and meeting abstracts and presentations. While the use of tau PET imaging rapidly increased in research and in clinical trials over the past few years, there is no standardization pipeline for the quantification of tau imaging across tau tracers and quantification software.
2. **Interpretation:** We built a global and several regional universal masks for the sampling of tau PET scans based on the most commonly used tau PET tracers. We then derived a universal scale across tracers, the CenTauR_z_, to measure the tau signal.
3. **Future directions:** Standardised quantification will facilitate the derivation of universal cut-off values, merging of large cohorts, and comparison of longitudinal changes across tracers and cohorts both in clinical studies and therapeutic trials.

## Background

Tau positron emission tomography (PET) imaging is the most recent addition to the arsenal of tools for the in vivo assessment of neurodegenerative proteinopathies. Prior to this development, the presence and extent of aggregated tau in the brain could only be characterized using postmortem examination [1]. Despite the challenges inherent to imaging tau pathology, which include its intracellular location, the presence of multiple human tau isoforms (three repeat (3R), four repeat (4R)), morphologies (paired helical filament (PHF), straight filament (SF), numerous post-translational modifications (e.g. phosphorylation, truncation, nitration), and, in the case of Alzheimer’s disease (AD), lower concentrations than amyloid-β (Aβ) in colocalizing tau and Aβ deposits (for review see [2, 3]) —there has been a tremendous amount of progress in the last few years, with several selective tau tracers identified and increasingly used for human imaging studies. These tracers have been shown to be largely specific for the mixed 3R/4R paired helical filament (PHF) tau pathology characteristic of AD and Down syndrome and have helped further our understanding of tauopathies as well as the relationship of between Aβ, tau, neurodegeneration and cognitive decline in AD [4-11].

In addition to the idiosyncratic characteristics of tau aggregates, and their asymmetric and heterogeneous brain distribution, a major obstacle to the widespread implementation of tau imaging in therapeutic trials or comparing the findings of investigational imaging studies across cohorts and institutions is that tau tracers differ in their molecular structures and display a range of tau binding affinities, in vivo kinetics, and degree of non-specific binding, as well as distinct regional patterns of “off-target” and non-specific binding. Such differences lead to disparities in PET-derived standardised uptake value ratios (SUVR) measurements between tracers, as highlighted by several head-to-head studies comparing different tau tracers [12, 13]. It is also important to note that most of these tau tracers do not reach apparent steady state in regions with high tau pathology during the scanning period, and while the use of semi-quantitative estimates such as SUVR was adopted early in the implementation of these tracers as a compromise to make PET imaging studies less burdensome to clinical populations, *a priori* kinetic modeling studies of tau tracers in early development stages may have led to further optimization of scanning protocols to be less biased to tau signal [14-17]. When added to the use of diverse quantitative approaches and different regions of interest, these methodologic differences conspire to decrease reproducibility and pose a challenge when trying to compare tau outcomes across cohorts or in therapeutic trials that use different tau tracers. A further obstacle within the tau PET field is the definition of a reliable, consistent and reproducible threshold of abnormality across tracers. One of the issues relates to the actual utility of a cut-off given the continuous nature of Aβ or tau deposition [18, 19]. While thresholds are arbitrary, in order to adopt one, it needs to be shown that it is relevant and accurate from a diagnostic and/or prognostic point of view [20, 21]. In essence, biomarker thresholds should be adopted for a specific purpose that is directly related to the clinical question under scrutiny. From a clinical perspective, a visual binary (positive/negative) status will help separate those subject with a significant aggregated protein burden in the brain that is likely to explain the clinical syndrome from those with a low pathologic burden that is likely to be clinically insignificant. Similar dilemmas arise in research settings.

In response to similar challenges faced earlier with Aβ PET [22], a standardization method was developed whereby Aβ PET outcome data acquired using different Aβ tracers and methods was normalized to a 100-point scale, the units of which were termed “Centiloids,” using a linear scaling procedure [22]. While the method transforms all Aβ tracers’ semiquantitative results into a single universal scale and because sampling was only based on ^11^C-PIB, the idiosyncratic binding properties of these Aβ tracers remain unaccounted for so they might be more or less sensitive or accurate for making a statement about a similar index of cerebral Aβ burden. Furthermore, while the pattern of Aβ deposition throughout the brain is relatively uniform across subjects, and thus a single universal target mask provides reproducible statements of Aβ in the brain, the deposition of tau, especially at the early stages, tends to be more heterogeneous[23], requiring a more regional approach to the sampling of target areas.

In the present study, we aimed to standardize tau PET results by establishing the location and amount of abnormality of tau aggregates in the brain, and expressing them in a universal standard scale, the units of which are termed “CenTauRs”—using tau PET data from the six most commonly used tracers (^18^F-floraucipir, ^18^F-MK6240, ^18^F-PI2620, ^18^F-PM-PBB3, ^18^F-RO948, and ^18^F-GTP1) and an approach similar to the one used in the Centiloid project.

## METHODS

This study involved 1,060 participants from various cohorts (AIBL, ADNI, BioFINDER), academic institutions (National Institutes for Quantum and Radiological Science and Technology, Chiba), as well as industry (Life Molecular Imaging, Genentech). All participants underwent a tau PET scan and a structural MRI (for complete details, see Supplementary Methods 1). All participants were assigned a diagnosis of cognitively unimpaired (CU), mild cognitive impairment (MCI) or AD dementia by the entity providing the data. Criteria for assigning participant diagnosis can be found elsewhere [15, 24-27]. Aβ status (Aβ+ or Aβ-) was defined using either Aβ PET or the Aβ42/Aβ40 ratio in cerebrospinal fluid (CSF). ANOVA was used to determine any significant demographic difference between cohorts.

### Image processing

Tau scans were spatially normalized using principal component analysis (PCA)-based Computational Analysis of PET by AIBL (CapAIBL) [28], which is a publicly available cloud based platform where PET images are spatially normalised to a standard template using an adaptive atlas approach (https://capaibl-milxcloud.csiro.au), and Statistical Parametric Mapping (SPM, version 8) using the standard pipeline for the Centiloid method described in Klunk et al. [22]. All spatially normalized scans were visually assessed to ensure proper registration, especially in the mesial temporal lobe (MTL) [29]. In the case of SPM, all scans that did not pass visual assessment were reprocessed using a different orientation matrix until they passed a visual quality check (QC). Scans that failed visual QC three times in a row were excluded from further analysis. In the CU group, Aβ-scans were excluded if the presence of tau was visually detected in the cortex or in the MTL. We defined a sub-cerebellar cortex region based on the Centiloid cerebellum cortex mask as reference region, excluding the upper part (slice > -37) of the cerebellum to avoid off-target binding often observed in the cerebellar vermis, and also the lower part (slice < -47) to avoid quantification challenges such as partial volume, low axial sensitivity, and out-of-field scatter (Supplementary Figure 1).

For each tracer and normalization approach (i.e., CapAIBL, SPM), we averaged all CU Aβ- and AD Aβ+ scans separately, generating mean CU Aβ- and AD Aβ+ images. We then subtracted the CU Aβ-mean image from the AD Aβ+ mean image to generate a difference image. After exploring several thresholds, the resultant difference-image was thresholded at 1/3 of the difference in the inferior temporal lobe. This threshold produced large and consistent VOIs across tracers of areas of the brain with the greatest tau load. We then constructed a “universal” tau mask from the intersection (i.e., spatial overlap) of the six tracer-specific masks. An MRI-derived grey matter mask obtained from the FreeSurfer segmentation of 100 MRIs (independent dataset) at PET resolution was then applied to the universal mask to only sample cortical regions. The resulting mask was then mirrored and fused to remove the hemispherical asymmetry of tau pathology. Lastly, an additional four sub-regions were defined within the constraints of the universal mask: Mesial Temporal, Meta Temporal, Temporo-Parietal and Frontal ROIs (Supplementary Methods 2). Agreement between masks was assessed using the Dice index, which is a measure of the similarity between various images. Finally, for each tracer, the mean and standard deviation of the CU Aβ-subjects were used to generate CenTauR z-scores in each of the five ROIs, similar to what was previously proposed by Vemuri and colleagues [30].

### Visual subtype classification

78 ^18^F -MK6240 AD Aβ+ scans from the AIBL cohort were visually were visually rated by two readers (CCR and NK), blind to participant characteristics, resulting in consensus visual reads, as previously described [31]. Briefly, scans were rated as i) *tau negative* (no tracer retention or minimal (unilateral or bilateral) entorhinal cortex retention; ii) *limbic predominant* (pronounced tracer retention in the MTL with no cortical retention); iii) *hippocampal sparing* (cortical tau tracer retention with no or minimal MTL signal); or iv) *typical* (MTL and cortical tracer retention).

## RESULTS

Participant characteristics by tau PET tracer are summarized in Supplementary Table 1. Overall, participants from the ^18^F-GTP1 and ^18^F-PM-PBB3 cohorts were significantly younger compared to participants from the other cohorts. Compared to the MCI and AD dementia groups, CU Aβ+ participants were significantly older (F-stat=3.9, p<0.002) and had fewer males (F-stat=3, p<0.005). No significant differences in age, gender, MMSE or CDR were found between the AD Aβ+ patients from the different cohorts (F-stat=1, p=0.4).

### Tau mask sampling

23 scans (eight ^18^F-RO948, one ^18^F-GTP1, five ^18^F-PI2620, one ^18^F-FTP, eight ^18^F-PM-PBB3) did not pass visual QC using the SPM pipeline or did not have an MRI of sufficient quality while only one scan did not pass visual QC when using both CapAIBL and SPM. A further six CU Aβ-were visually excluded due to the presence of tracer uptake in the MTL. These 29 scans were excluded from further analysis.

SPM tracer-specific masks showed a reasonable overlap, with a global Dice score of 0.58 [95% CI, 0.52-0.61] and a Dice score in the cortical mask of 0.61 [95% CI, 0.60-0.69]. All masks included the mesial temporal, meta-temporal, posterior cingulate/precuneus and sub frontal regions. The CenTauR mask overlaid on an MRI template is shown in Figure 1, while the subregion masks are shown in Supplementary Figure 2. None of the known off-target signal regions were discernible in the five masks (Supplementary Figure 3).

**Figure 1:**
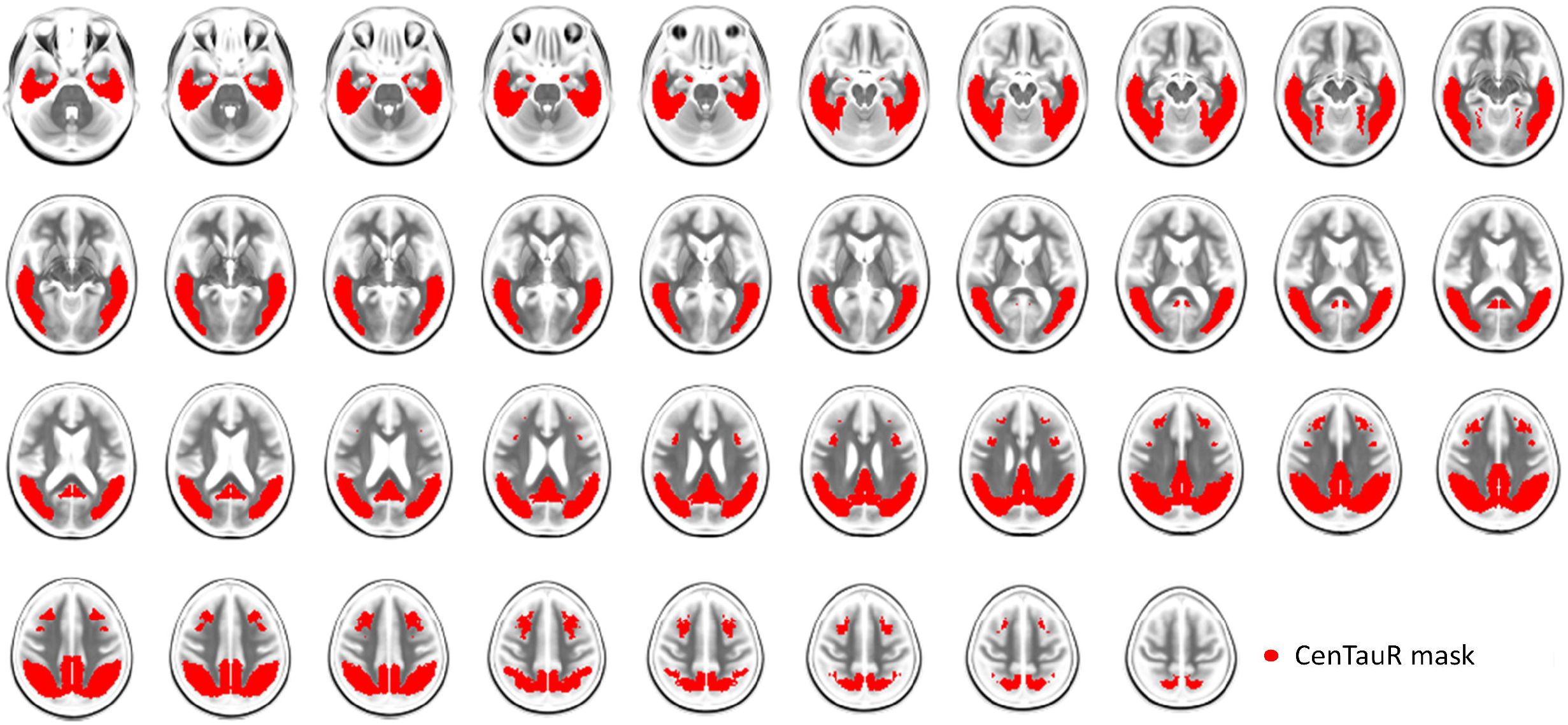
CenTauR mask overlaid on an MRI template

Both quantitative pipelines provided very similar tau masks, with a Dice score of 0.75 between universal masks generated using CapAIBL and SPM. Part of this difference was due to the CapAIBL mask not being in MNI space, which required resampling to be compared to the SPM mask. In the remainder of this paper, we only use the masks defined using the SPM pipeline.

### CenTauR_z_ (CTR_z_) quantificationError! Reference source not found

1 provides the regional equations to convert SPM-based SUVR values into CTR_z_ for each of the six tau tracers included in the study. Figure 2 displays the box plot of the Meta Temporal CTR_z_ for CU Aβ- and AD Aβ+ individuals. CTR_z_ for the other four ROIs are presented in Supplementary Figure 5 and CapAIBL CTR_z_ are displayed in Supplementary Figure 6. Using a threshold of 2 CTR_z_ in the Meta Temporal ROI, all tracers showed high discriminative accuracy for the separation of AD Aβ+ from CU Aβ-individuals (accuracy=0.96 [min=0.95-max=1], sensitivity=0.91 [0.78-1], specificity=0.97 [0.93-1]) with mean CTR_z_ scores for the six different AD cohorts ranging from 8.1 to 22 (Figure 2 and Supplementary Figure 4). Similar accuracies were observed using the Mesial Temporal (accuracy=0.95 [0.90-1], sensitivity=0.90 [0.83-1], specificity=0.97 [0.95-1]) and Temporo-Parietal (accuracy=0.94 [0.90-1], sensitivity=0.88 [0.76-1], specificity=0.96 [0.95-1]) ROIs, while the accuracy for the Frontal ROI (accuracy=0.91 [0.81-1]) was somewhat lower due to lower sensitivity (sensitivity = 0.73 [0.5-1]); whereas specificity (specificity = 0.97 [0.91,1]) was similar to that for the Meta Temporal ROI.

**Figure 2:**
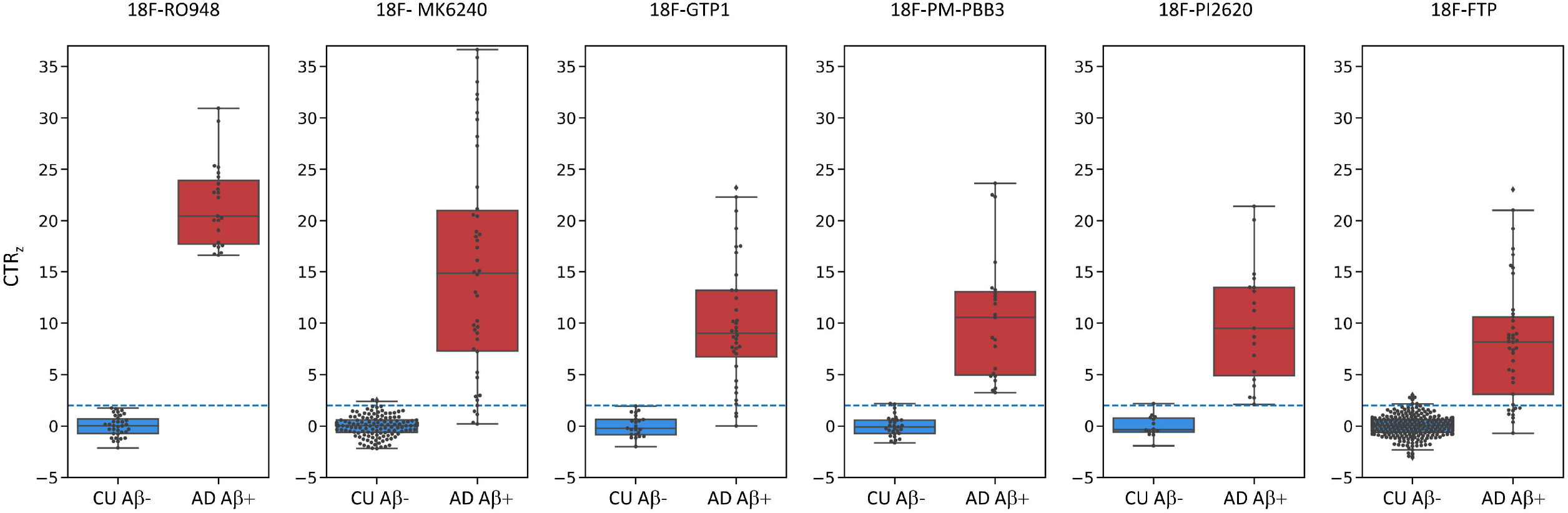
Comparisons of the CenTauR_z_ (CTR_z_) in the Meta Temporal ROI between CU Aβ- and AD Aβ+ for the 6 tau tracers. The blue dashed line corresponds to 2 CTR_z_.

Figure 3 shows boxplots of CTR_z_ scores in the 5 different ROIs. The AD Aβ+ group had significantly higher CTR_z_ scores across ROIs compared to all other cognitive groups (Welch’s T>7.6). CU Aβ+ had significantly higher CTR_z_ compared to CU Aβ- and MCI Aβ-in all regions with the strongest effect size in the Mesial Temporal ROI (Welch’s T>6) and the lowest in the Frontal ROI (Welch’s T∼3). Among the CU Aβ+, 36% had a CTR_z_ higher than 2 in the Mesial Temporal, 29% in the Meta Temporal, 21% in the Temporo-Parietal, 12% in the Frontal and 23% in the global, while these prevalences were respectively 77%, 63%, 58%, 41%, 60% for the MCI Aβ+ group, and 91%, 90%, 87%, 73%, and 88% for the AD Aβ+ and around 4% and 2.5% in all regions for the MCI Aβ- and CU Aβ-respectively.

**Figure 3:**
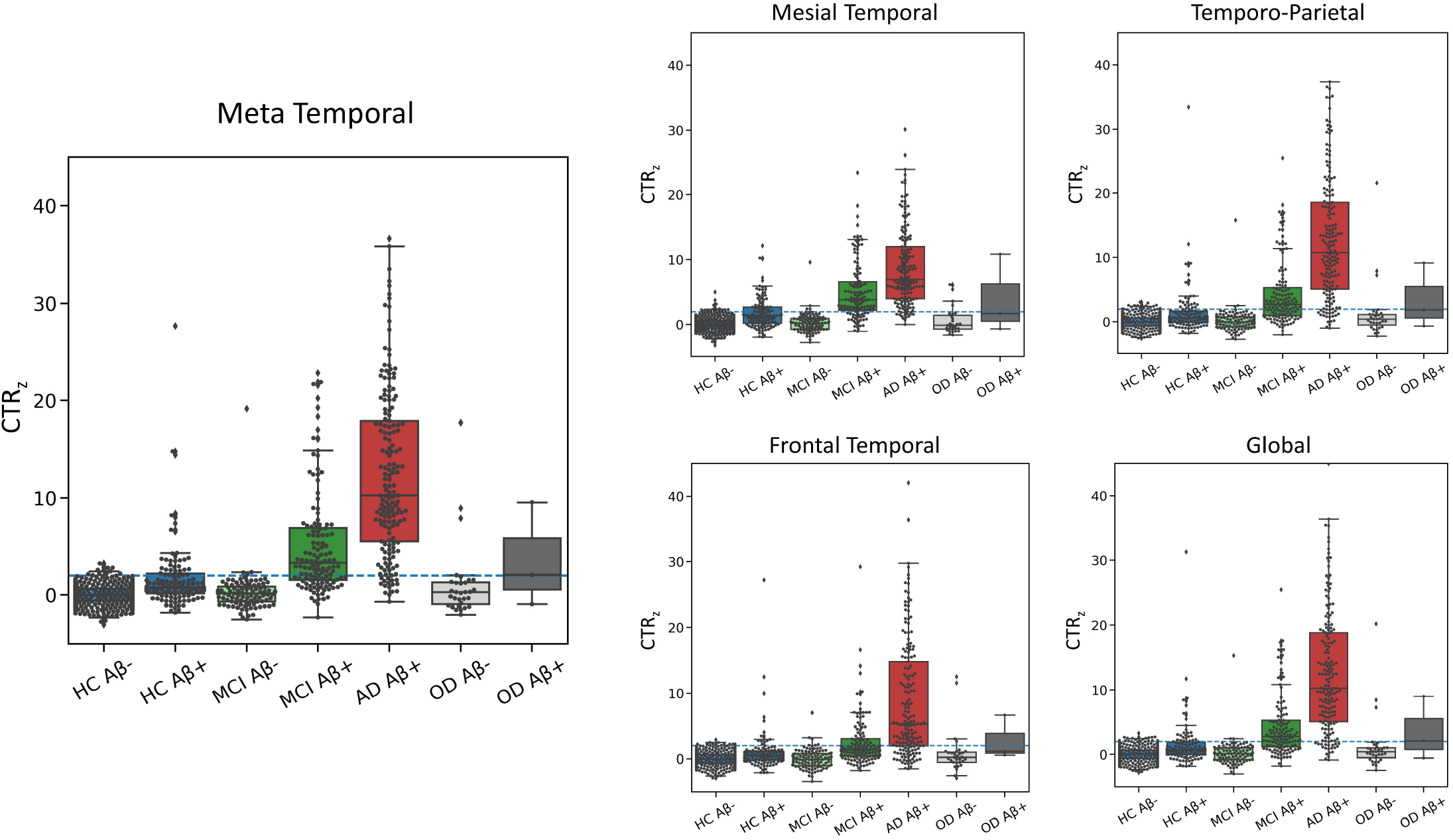
Boxplots of the ROI CTR_z_ in the different ROI. The blue dashed line corresponds to 2 CTR_z_.

### CapAIBL versus SPM pipeline

The equations to convert CapAIBL SUVR values into CTR_z_ scores are presented in Supplementary Table 2. Converting slopes between CapAIBL and SPM were of the same rank order except for the Temporo-Parietal and Frontal ROIs for ^18^F-PM-PBB3, due to the slightly higher standard deviation of the CapAIBL SUVRs in the CU Aβ-group. The correlation between CTR_z_ scores from SPM and CapAIBL was 0.99 in the Meta Temporal ROI, 0.98 in the Mesial, Temporo-Parietal and Global ROIs, and 0.89 in the Frontal ROI (Figure 4). Using an arbitrary threshold of 2.0 CapAIBL CTR_z_ in the Meta Temporal region, all tracers showed high discriminative accuracy for the separation of AD Aβ+ from CU Aβ-individuals (accuracy=0.95 [0.93-1], sensitivity=0.89 [0.78-1], specificity=0.98 [0.96-1]), with mean CTR_z_ for the different AD cohorts ranging from 7.6 to 20.6 (Supplementary Figures 6, 7 and 8).

**Figure 4:**
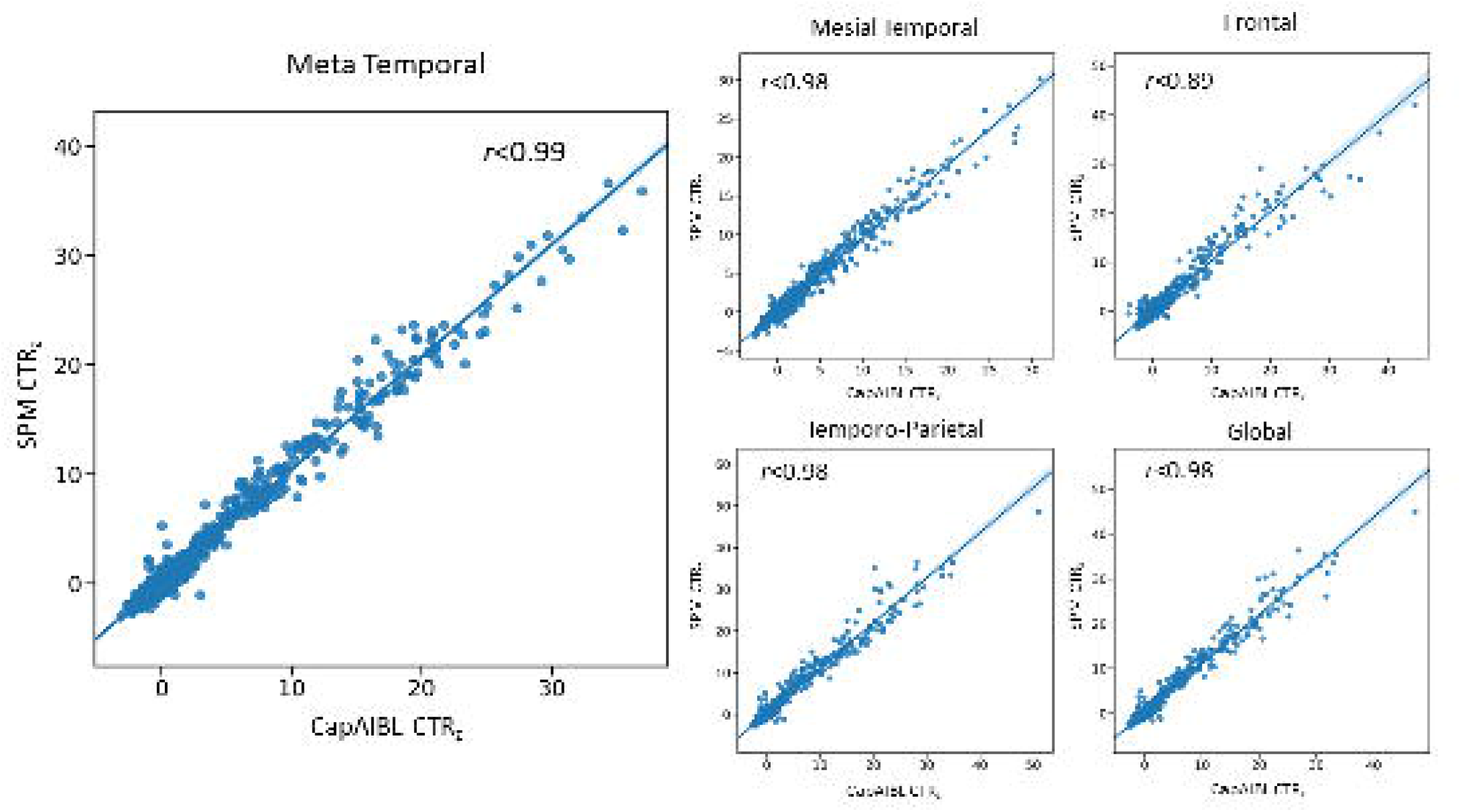
Comparison of the CTR_z_ generated with SPM (y-axis) and with CapAIBL (x-axis)

Supplementary Figures 9 and 10 display the association between CTR_z_ scores and Aβ+ PET Centiloid values across the five ROIs. As previously reported [32], individuals with a Centiloid value below 50 and a CTR_z_ value above 2 in the Meta Temporal or Temporo-Parietal ROIs were rare but became increasingly more common as Centiloid values increased. Similarly, very few individuals with a Frontal CTR_z_ values above 2 had Centiloid values below 70. Scatter plots showing the relationship between CapAIBL derived CTR_z_ scores and Centiloids are shown in Supplementary Figure 10.

Scatter plots showing CTR_z_ scores in the Meta Temporal and Temporo-Parietal ROIs as a function of CTR_z_ scores in the Mesial temporal for ^18^F-MK6240 are presented in Figure 5. Visual classifications (i.e., tau negative, limbic predominant, hippocampal sparing and typical) are colour coded. A CTR_z_ > 2 in the Mesial Temporal ROI accurately differentiated tau negative scans from all other classifications (accuracy=0.92, sensitivity=0.97, specificity=0.60); applying a threshold of 2 CTR_z_ in the Mesial Temporal and in the Meta Temporal together slightly increased the accuracy of detecting tau negative scans (accuracy=0.94, sensitivity =1.0, specificity =0.60). Using CTR_z_ > 2 in Mesial Temporal ROI and <2 in the Meta Temporal ROI, yielded an accuracy of 0.92 to detect Limbic predominant individuals. Using CapAIBL the specificities and accuracies were slightly improved (Tau negative: accuracy=0.95, sensitivity=0.97, specificity=0.80; Limbic predominant: accuracy=0.92, sensitivity=0.96, specificity=0.57, Supplementary Figure 11 & 12).

**Figure 5.**
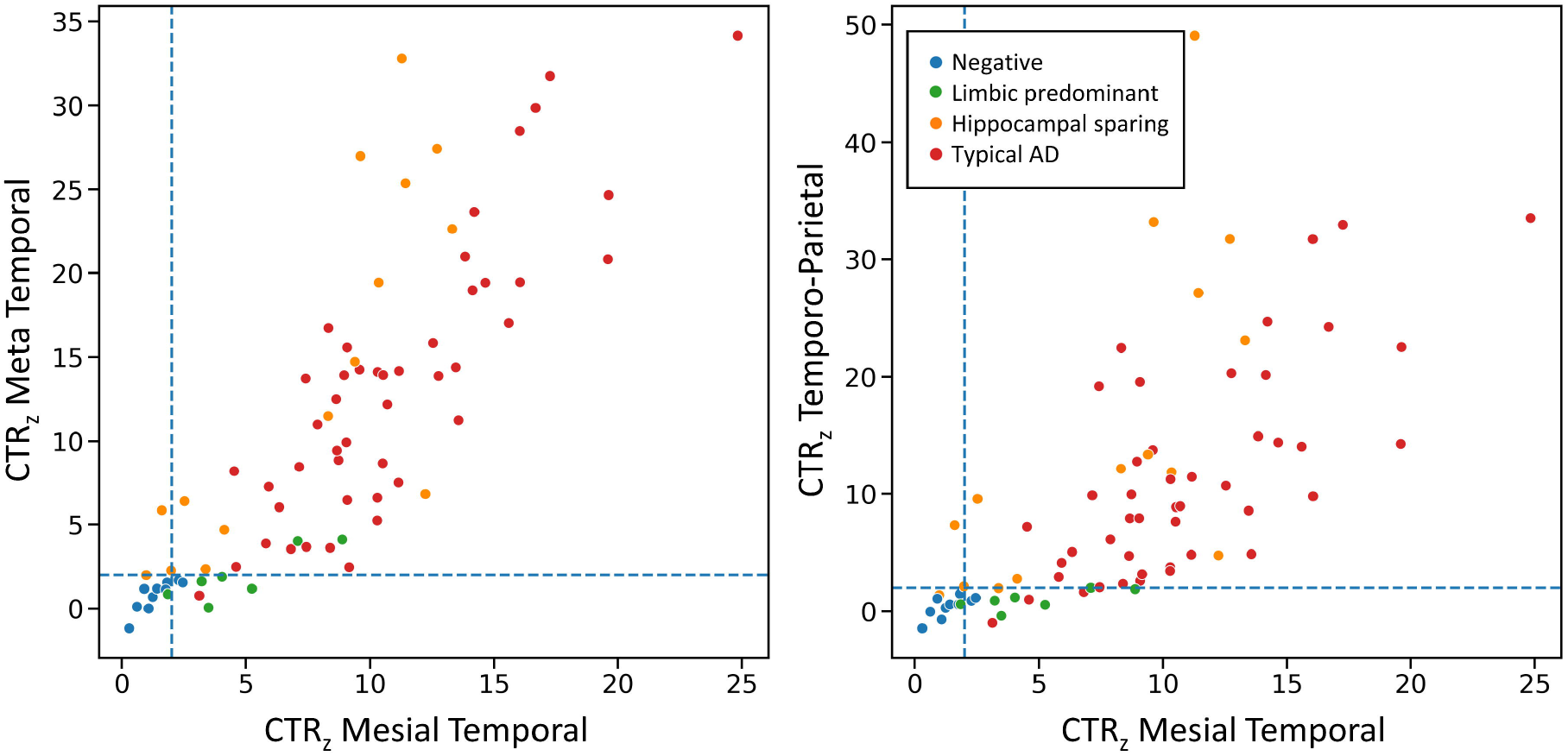
Scatter plots of the CTR_z_ in the Meta Temporal and temporo-Parietal as a function of the CTR_z_ in the Mesial temporal from the ^18^F-MK6240 AIBL cohort. Points are coloured depending on their visual reads. The blue dashed lines correspond to 2 CTR_z_.

## DISCUSSION

In the present work we described the CenTauR_z_ scale, a method that facilitates the expression of the level of abnormality of the semiquantitative tau PET signal at both a regional and global level. Also, the CenTauR_z_ scale allows, by incorporating the intrinsic “noise” of each tau tracer into the measurement, the generation of a universal scale of tau pathologic burden across tracers. The two pipelines used to quantify brain PET imaging (CapAIBL and SPM) generated consistent results in quantifying tau scans in all ROIs, with high discriminative power in distinguishing AD Aβ+ from CU Aβ- and tau negative scans from limbic predominant, hippocampal sparing and typical AD tau scans when using a threshold of > 2 CTR_z_ in different ROIs.

An important aspect, both for clinical interpretation and for therapeutic trials, is the selection of brain regions sampled in order to capture the distribution of tau, how this index of tau load changes over time, and what CTR_z_ level is considered high tau [33]. Given the low spatial resolution of PET, it can be counterproductive to impose a neuropathological piecemeal staging system, such as those proposed by Braak and Braak [34] or Delacourte [35], to the sampling of tau PET images [36, 37]. Atypical and heterogeneous presentations of tau deposits, and how they intimately relate to the clinical phenotype [34, 35], are missed by the incrementally sequential Braak staging. Applying the Braak or Delacourte staging [34, 35] is further complicated by the different neuropathological subtypes of tau deposition in AD [38]. From the pathological AD subtypes, only the typical (reported to be between 55-75% in different series) [39-41] completely fulfills the sequential Braak stages. Several reports have shown that a meta-temporal region [42], or a temporoparietal (including posterior cingulate) AD-signature region [43, 44] outperforms the Braak staging for the early detection of cortical tau, for establishing the differential diagnosis of AD vs non-AD neurodegenerative conditions [45], as well as for capturing longitudinal changes in cortical tau signal. These regions seem to perform reliably across different tau tracers and use sites and, despite these tracers presenting different dynamic ranges, they yielded the same cut-off for abnormality in different cohorts [46]. While the use of tau imaging for disease staging is strongly recommended [47], the use of neuropathological staging should be applied carefully, not as an *a priori* condition, but as the result of the actual observed pattern of tau deposition on the PET images. Furthermore, it has been shown that tau imaging, at least with ^18^F-FTP [48], can reliably detect a B3 stage (equivalent to Braak V-VI), so attempting to classify earlier Braak stages using this tracer, with its high level of non-specific binding [49], would likely yield less reliable results. Similar issues may apply to other tau tracers. Such considerations argue against using current neuropathological staging approaches, especially because it progresses from very small regions (Braak I-II) that are susceptible to partial volume effects and easily contaminated by off-target binding, to very large regions (Braak V-VI) that encompass large portions of the cerebral cortex and subcortical structures, making it impractical for implementation in clinical studies, and foremost, in therapeutic trials. Our method is designed to capture tau levels and distribution in the brain as well as tau progression and most of the reported heterogeneities in tau PET studies, such as primary age-related tauopathy (PART) and proposed subtypes and heterogeneity in the patterns of tau distribution [31] [50]. Similar methods can be used to the brain region selected as reference to scale the tissue ratios. Attempts to define a universal cerebellar tau mask are already underway (*GOGOLA, MINHAS in this issue*), but will require testing with all tau tracers to assess whether it improves the CTR_z_ accuracy.

There are several limitations of the present study. Firstly, similar to the Centiloid method, the mask and scales for some of the tau tracers included were generated from a limited number of available participant datasets. Secondly, the masks and scales were generated with elderly CU Aβ-controls and AD Aβ+ patients. A scale generated with young adult controls devoid of cortical tau pathology might hypothetically prove more sensitive to low levels of tau pathology. That said, ongoing studies with ^18^F-MK6240 and ^18^F-FTP comparing young adult controls with elderly controls show no significant differences in the tau signal [51] between young and elderly controls. Thirdly, the performance of the masks and scales were not tested in longitudinal studies and therefore we cannot assess the reproducibility of the method. However, the CenTauR framework is flexible in several key aspects: a) while the results presented here are the average of left and right hemispheres, data can be expressed unilaterally to characterize potential asymmetries in tau deposition; b) in order to capture early cortical tau deposition in the inferior and middle temporal gyri, the MTL CTR_z_ could be subtracted from the meta temporal CTR_z_; c) similar to what was proposed with the Centiloid method, it allows to resample a CTR_z_ parametric image, either with a different atlas template, employing SPM or with a different image analysis pipeline or software, once all voxels are transformed into CTR_z_ parametric images using one of the provided equations (for a global transformation, we suggest using the temporoparietal equation (Supplementary Figure 12); and d) it provides a comprehensive scheme to facilitate and standardize head-to-head comparisons between tau tracers [52, 53]. Lastly, the modular approach also allows the examination of certain brain regions separately given that they behave differently over time, with for example the MTL accumulating tau early but also plateauing early, or the temporoparietal that seems to be the most sensitive region to capture tau accumulation in the brain, and likely large enough to provide robust statements of changes in tau burden in a clinical trial.

In conclusion, we constructed several universal tau PET specific cortical masks for the AD continuum based on all the commonly used tau tracers, and a universal standard scale, the CenTauR_z_, designed to capture the location and degree of abnormality of tau pathology that can be applied across tracers and across centres. While the CenTauR scheme does not answer *all* questions about measuring tau deposits, it establishes a robust and reproducible standard framework from which to build upon, and to be implemented in the clinic and applied in therapeutic trials.

**Table 1:** Conversion equations from SPM SUVR to CTR_z_

| Tracer | Global | Mesial<br>Temporal | Meta Temporal | Temporo<br>Parietal | Frontal |
| --- | --- | --- | --- | --- | --- |
| <sup>18</sup>F-RO948 | 13.05 x – 15.57 | 11.76 x – 13.08 | 13.16 x – 16.19 | 13.05 x – 15.62 | 12.61 x – 13.45 |
| <sup>18</sup>F-FTP | 13.63 x – 15.85 | 10.42 x – 12.11 | 12.95 x – 15.37 | 13.75 x – 15.92 | 11.61 x – 13.01 |
| <sup>18</sup>F-MK6240 | 10.08 x – 10.06 | 7.28 x – 7.01 | 9.36 x – 10.6 | 9.98 x – 10.15 | 10.05 x – 8.91 |
| <sup>18</sup>F-GTP1 | 10.67 x – 11.92 | 7.88 x – 8.75 | 9.60 x – 11.10 | 10.84 x – 12.27 | 9.41 x – 9.71 |
| <sup>18</sup>F-PM-PBB3 | 16.73 x – 15.34 | 7.97 x – 7.83 | 11.78 x – 11.21 | 16.16 x – 14.68 | 15.7 x – 13.18 |
| <sup>18</sup>F-PI2620 | 8.45 x – 9.61 | 6.03 x – 6.83 | 7.78 x – 9.33 | 8.21 x – 9.52 | 9.07 x – 9.01 |

## Data Availability

All data produced in the present study are available upon reasonable request to the authors.

## Declarations

### Consent Statement

All participants gave written consent for publication of de-identified data.

## Footnotes

1 The CenTauR masks are freely available at https://www.gaain.org/to-be-defined

