## Supplementary material for "CenTauR: Towards a Universal Scale and Masks for Standardizing Tau Imaging Studies"

**Supplementary Methods 1. Tau PET**

**^18^F-MK6420 (AIBL cohort)**

From the AIBL cohort, 189 CU (157 Aβ- and 32 Aβ+), 57 MCI (22 Aβ-, 35 Aβ+) and 42 AD Aβ+ and 12 other dementia (OD) Aβ- underwent a 20-minute ^18^F-MK6240 scan, acquired 90 minutes post-injection of 185 MBq (+/- 10%) and a 50-70min ^18^F-NAV4694 (200 MBq). All radiotracers were synthesized in-house at Austin Health, Melbourne, Australia. PET scans were acquired on one of two scanners: Philips TF64 PET/CT or a Siemens Biograph mCT. A low dose CT was obtained for attenuation correction.

**^18^F-Flortaucipir (AIBL and ADNI cohorts)**

54 CU (37 Aβ-, 17 Aβ+), 10 MCI (4 Aβ-, 6 Aβ+) and 18 AD Aβ+ from the AIBL cohort underwent a 20-minute ^18^F-Flortaucipir scan, acquired 80 minutes post-injection of 240 MBq (+/- 10%) and a 50-70min ^18^F-Florbetapir (360 MBq +/- 10%). Both radiotracers were synthesized in-house at Austin Health, Melbourne, Australia. PET scans were acquired on one of two scanners: Philips TF64 PET/CT or a Siemens Biograph mCT. A low dose CT was obtained for attenuation correction.

We also included 186 CU Aβ-, 60 CU Aβ+, 70 MCI Aβ-, 65 MCI Aβ+, 32 AD Aβ+, 8 OD Aβ- from the Alzheimer's Disease Neuroimaging Initiative (ADNI) database (adni.loni.usc.edu)^[1]^.

**^18^F-PI2620 (Life Molecular Imaging)**

16 CU (14 Aβ-), 19 AD Aβ+ and 12 OD were administered a single dose of ^18^F-PI2620 (338.7±20.9 MBq; range, 262.7–359.8 MBq) through a venous catheter followed by a 10-mL saline flush. Scans were acquired between 70 to 90min post injection on a Siemens ECAT EXACT HR1 camera and Philips Gemini TF64.

β-amyloid PET images were acquired according to the prescribing information using the same PET scanner with ^18^F-NAV4964 and ^18^F-Florbetaben tracers. A T1-weighted MRI scan was acquired as part of the screening on a Siemens Espree 1.5-T or on a 3T Skyra scanner.

**^18^F-RO948 (Biofinder cohort)**

From the BioFINDER2 cohort 34 CU Aβ- and 23 AD Aβ+ underwent a 20 minute ^18^F-RO948 scans acquired 70 min post injection of 365 ± 20 MBq of ^18^F-RO948 on a digital GE Discovery MI scanner (General Electric Medical Systems). Low-dose CT scans were performed immediately prior to the PET scans for attenuation correction. Aβ status of the participants was determined using a threshold of 0.089 on the cerebrospinal fluid (CSF) Aβ42/Aβ40 ratio, as defined in clinical practice at the Sahlgrenska University Hospital, Mölndal, Sweden.

**^18^F-GTP1 (Genentech)**

23 CU (12 CU Aβ- and 11 CU Aβ+), 27 MCI (26 Aβ+) and 37 AD Aβ+ were scanned for tau using the ^18^F-GTP1 tracer.

^18^F-GTP1 images were acquired over a 30-minute window 60 minutes after injection after a mean (SD) bolus injection of 343 ± 31 MBq on Siemens HR+ PET or Biograph 6 PET-CT cameras at Invicro (New Haven, CT). Images were reconstructed with an iterative reconstruction algorithm (OSEM 4 iterations, 16 subsets) and a post hoc 5 mm Gaussian filter.

Each participant also underwent 20 minutes of serial PET imaging starting 50 minutes (±5 minutes) after injection of ^18^F-Florbetapir. Aβ PET were quantified in Centiloid.

**^18^F-PM-PBB3 (Chiba cohort)**

27 CU Aβ- and 24 AD Aβ+ underwent a tau PET scan for 20-min, 90 min after injections of ^18^F-PM-PBB3 (189.5 ± 22.5 MBq). 11C-PiB (injected dose: 521.2 ± 87.3 MBq) PET scan were also conducted with a 20-min acquisition 50 min after injections. PET assays were conducted with a Biograph mCT flow system (Siemens Healthcare), a ECAT EXACT HR+ scanner (CTI PET Systems, Inc.) was also utilized for 11C-PiB alternatively. The intrinsic spatial resolution was 5.9 mm in-plane and 5.5 mm full-width at half-maximum axially. Images were reconstructed using a filtered back projection algorithm with a Hanning filter (4.0 mm full-width at half-maximum).

All MR images were acquired with a 3-T scanner, MAGNETOM Verio (Siemens Healthcare).

**Supplementary Methods 2. Sub-region masks**

A Mesial Temporal, Meta Temporal, Temporo-Parietal and Frontal ROIs were respectively composed of the following derived FreeSurfer regions: the Mesial Temporal: entorhinal, parahippocampus and amygdala; the Meta Temporal region: entorhinal, parahippocampus, amygdala, fusiform, inferior and middle temporal gyri; the Temporo-Parietal: bankssts, cuneus, inferior-superior parietal, inferior-middle-superior temporal, istmuscingulate, lateral occipital, lingal, posterior cingulate, precuneus and superior marginal; the Frontal: caudate middle frontal, precentral, rostral middle frontal, Superior frontal.

**Supplementary Table 1:** Demographics of the different sub-cohorts

| Tracer | Subgroup | Sample size | Age | Gender  (% Male) | MMSE | CDR | Centiloid |
| --- | --- | --- | --- | --- | --- | --- | --- |
| ^18^F-MK6240 | CU Aβ- | 157 | 73.9 (4.9) | 47.1 | 28.7 (1.2) | 0.0 (0.1) | 3 (8) |
|  | CU Aβ+ | 32 | 76.1 (5.9) | 31.2 | 28.1 (1.7) | 0.1 (0.2) | 87 (42) |
|  | MCI Aβ- | 22 | 70.5 (7.9) | 45.5 | 27.6 (2.0) | 0.5 (0.1) | 2 (7) |
|  | MCI Aβ+ | 35 | 74.7 (6.8) | 60.0 | 25.9 (2.2) | 0.5 (0.0) | 116 (39) |
|  | AD Aβ+ | 42 | 71.4 (7.9) | 50.0 | 21.6 (4.8) | 0.9 (0.6) | 110 (43) |
|  | OD | 12 | 71.1 (5.4) | 58.3 | 23.3 (2.3) | 0.7 (0.3) | 12 (31) |
| ^18^F-FTP | CU Aβ- | 223 | 70.5 (6.1) | 37.3 | 29.0 (1.2) | 0.0 (0.1) | 1 (10) |
|  | CU Aβ+ | 77 | 74.4 (7.2) | 41.8 | 28.7 (1.3) | 0.0 (0.1) | 60 (27) |
|  | MCI Aβ- | 74 | 72.0 (8.2) | 61.8 | 28.2 (2.0) | 0.5 (0.1) | -2 (10) |
|  | MCI Aβ+ | 71 | 72.6 (6.8) | 50.9 | 27.1 (2.3) | 0.5 (0.1) | 77 (31) |
|  | AD Aβ+ | 50 | 74.0 (8.0) | 56.5 | 22.1 (3.9) | 0.8 (0.3) | 96 (32) |
|  | OD | 8 | 75.8 (7.7) | 83.3 | 23.5 (1.3) | 0.9 (0.5) | 5 (9) |
| ^18^F-PI2620 | CU Aβ- | 16 | 71.2 (7.5) | 14.3 | 25.6 (7.8) | 0.7 (1.2) | nan (nan) |
|  | CU Aβ+ | 2 | 70.0 (1.4) | 50.0 | 30.0 (0.0) | 0.2 (0.4) | nan (nan) |
|  | AD Aβ+ | 19 | 70.5 (7.2) | 25.0 | 20.4 (4.2) | 0.8 (0.3) | nan (nan) |
|  | OD | 12 | 67.1 (5.1) | 53.8 | 25.8 (3.3) | 0.5 (0.1) | nan (nan) |
| ^18^F-RO948 | CU Aβ- | 34 | 71.5 (3.8) | 44.1 | nan (nan) | nan (nan) | nan (nan) |
|  | AD Aβ+ | 23 | 71.4 (6.9) | 54.5 | nan (nan) | nan (nan) | nan (nan) |
| ^18^F-GTP1 | CU Aβ- | 12 | 63.1 (7.2) | 33.0 | 29.4 (0.9) | 0.0 (0.0) | 3 (14) |
|  | CU Aβ+ | 11 | 69.5 (3.0) | 36.4 | 29.3 (0.8) | 0.0 (0.0) | 57 (36) |
|  | MCI Aβ- | 1 | 71 | 0.0 | 30.0 | 0.5 | 12 |
|  | MCI Aβ+ | 26 | 69.2 (7.8) | 40.0 | 28.0 (1.4) | 0.5 (0.0) | 80 (31) |
|  | AD Aβ+ | 37 | 70.1 (7.0) | 58.3 | 21.5 (4.7) | 0.8 (0.4) | 82 (33) |
| ^18^F-PM-PBB3 | CU Aβ- | 27 | 64.5 (10) | 53.6 | 28.5 (1.6) | nan (nan) | nan (nan) |
|  | AD Aβ+ | 24 | 70.1 (11) | 52.4 | 21.7 (3.0) | nan (nan) | nan (nan) |

Abbreviations: Cognitively unimpaired (CU), Mild Cognitive impairment (MCI), Alzheimer’s disease (AD), other Dementia (OD).

**Supplementary Table 2:** Equation to convert CapAIBL SUVR to CTR_z_

| **Tracer** | **Mesial** | **Meta** | **Temporo-Parietal** | **Frontal** | **Global** |
| --- | --- | --- | --- | --- | --- |
| **^18^F-RO948** | 14.31 **x** - 15.86 | 13.27***x** - 16.23 | 13.63 **x** – 16.26 | 11.63 **x** - 12.17 | 13.67 **x** - 16.27 |
| **^18^F-FTP** | 9.93 **x** - 11.37 | 12.19 **x** – 14.54 | 13.08 **x** - 15.05 | 9.24 **x** - 9.88 | 12.62 **x** – 14.6 |
| **^18^F-MK6240** | 7.24 **x** - 7.15 | 8.74 **x** – 9.43 | 8.96 **x** – 9.31 | 9.55 **x** - 8.69 | 9.29 **x** – 9.32 |
| **^18^F-GTP1** | 7.35 **x** – 8.81 | 11.1 **x** – 12.39 | 13.81 **x** – 15.13 | 13.81 **x** – 15.13 | 13.17 **x** – 12.92 |
| **^18^F-PM-PBB3** | 6.90 **x** - 6.99 | 10.02 **x** - 9.64 | 12.08 **x** – 10.76 | 10.32 **x** – 10.76 | 12.16 **x** – 11.04 |
| **^18^F-PI2620** | 8.07 **x** – 8.83 | 8.81 **x** – 10.46 | 8.33 **x** – 9.70 | 11.07 **x** – 10.91 | 9.91 **x** - 11.09 |

**Supplementary Figure 1.** Reference region used in this analysis.

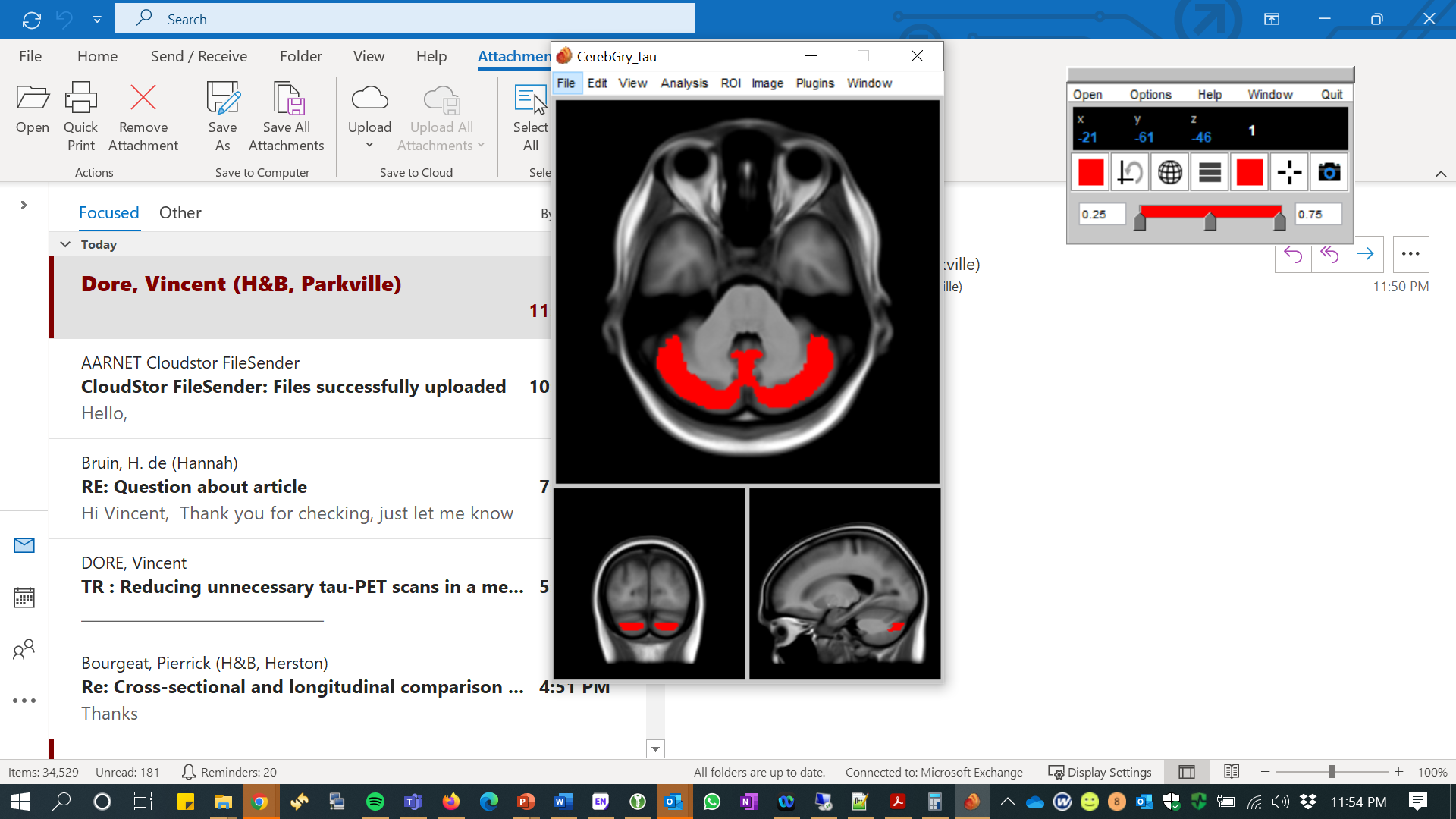

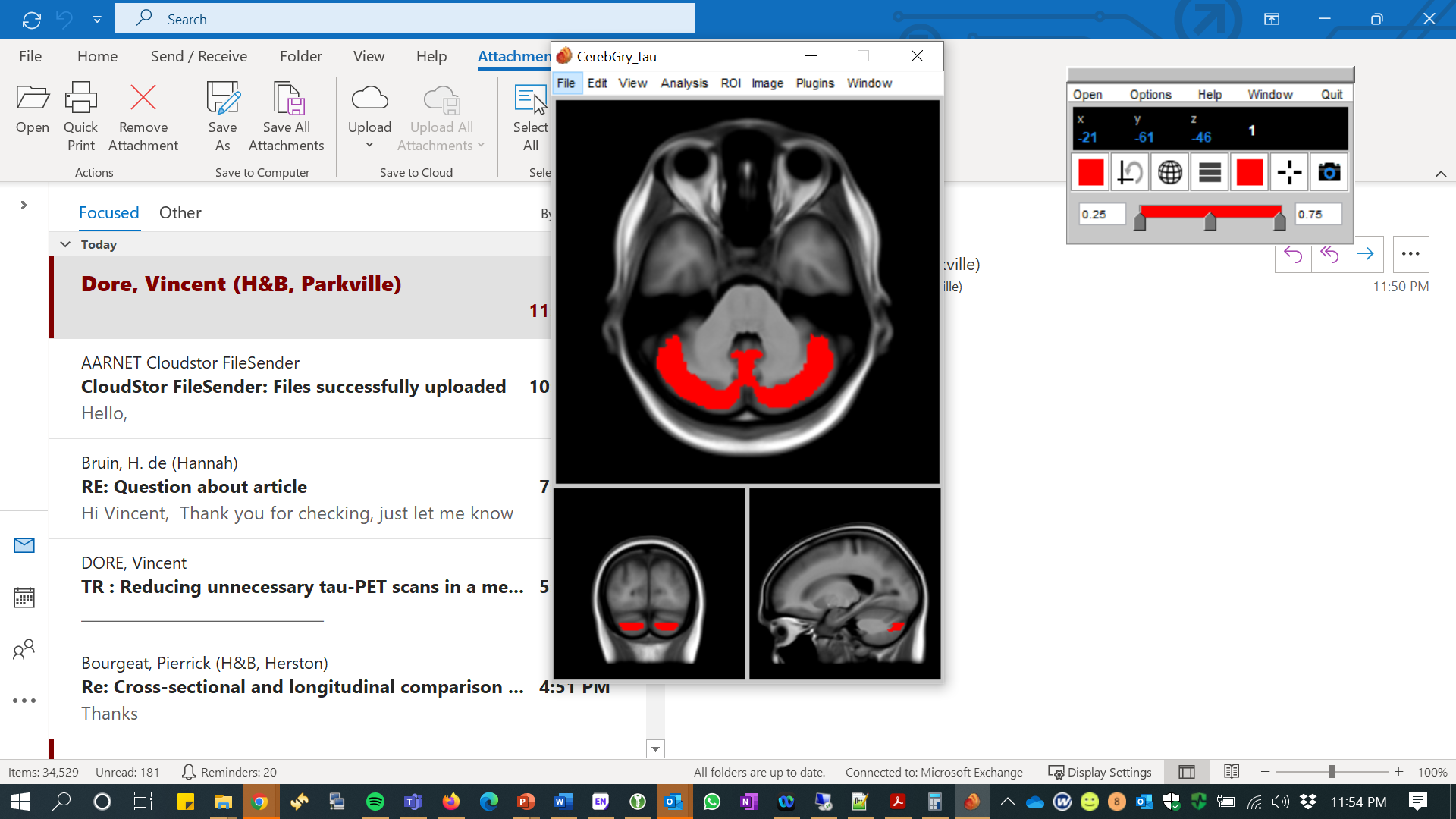

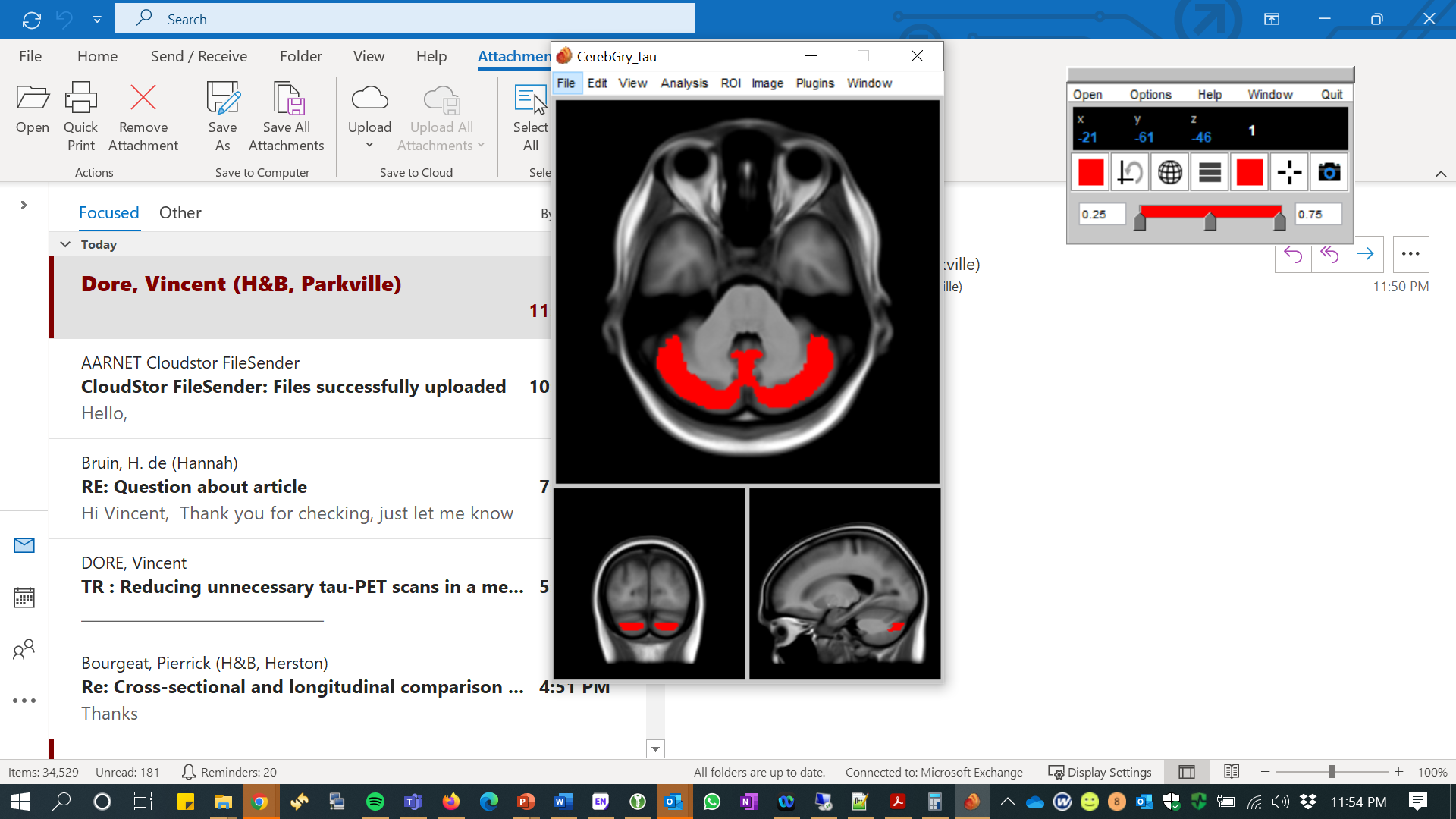

**Supplementary Figure 2:** CenTauR sub-cortical masks overlaid on an MRI template

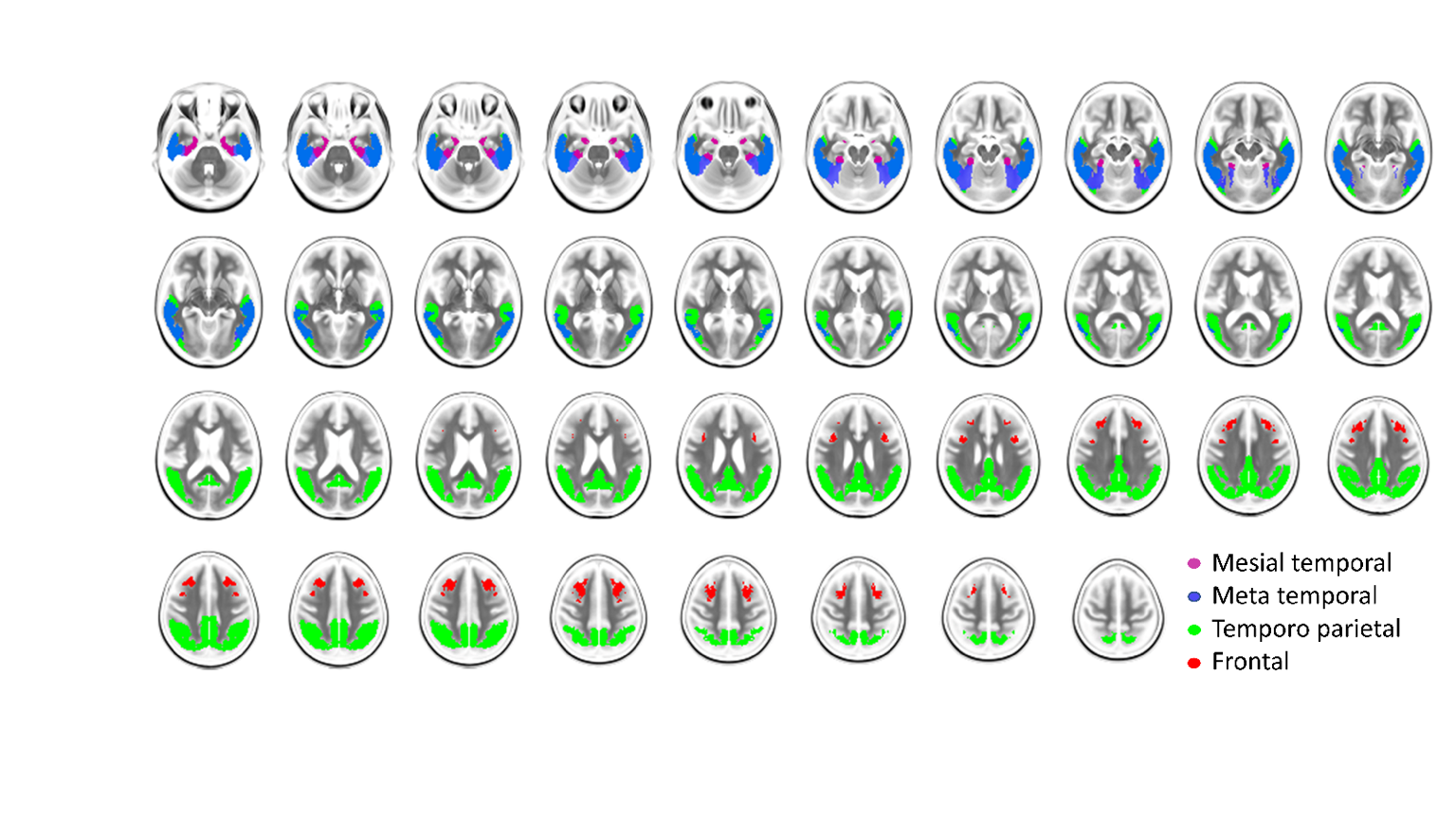

**Supplementary Figure 3:** Top two row, Flortaucipir with choroid plexus off-target binding, bottom row ^18^F-MK6240 scan with meninges uptakes. In red, superimposed is the CenTauR Meta temporal mask

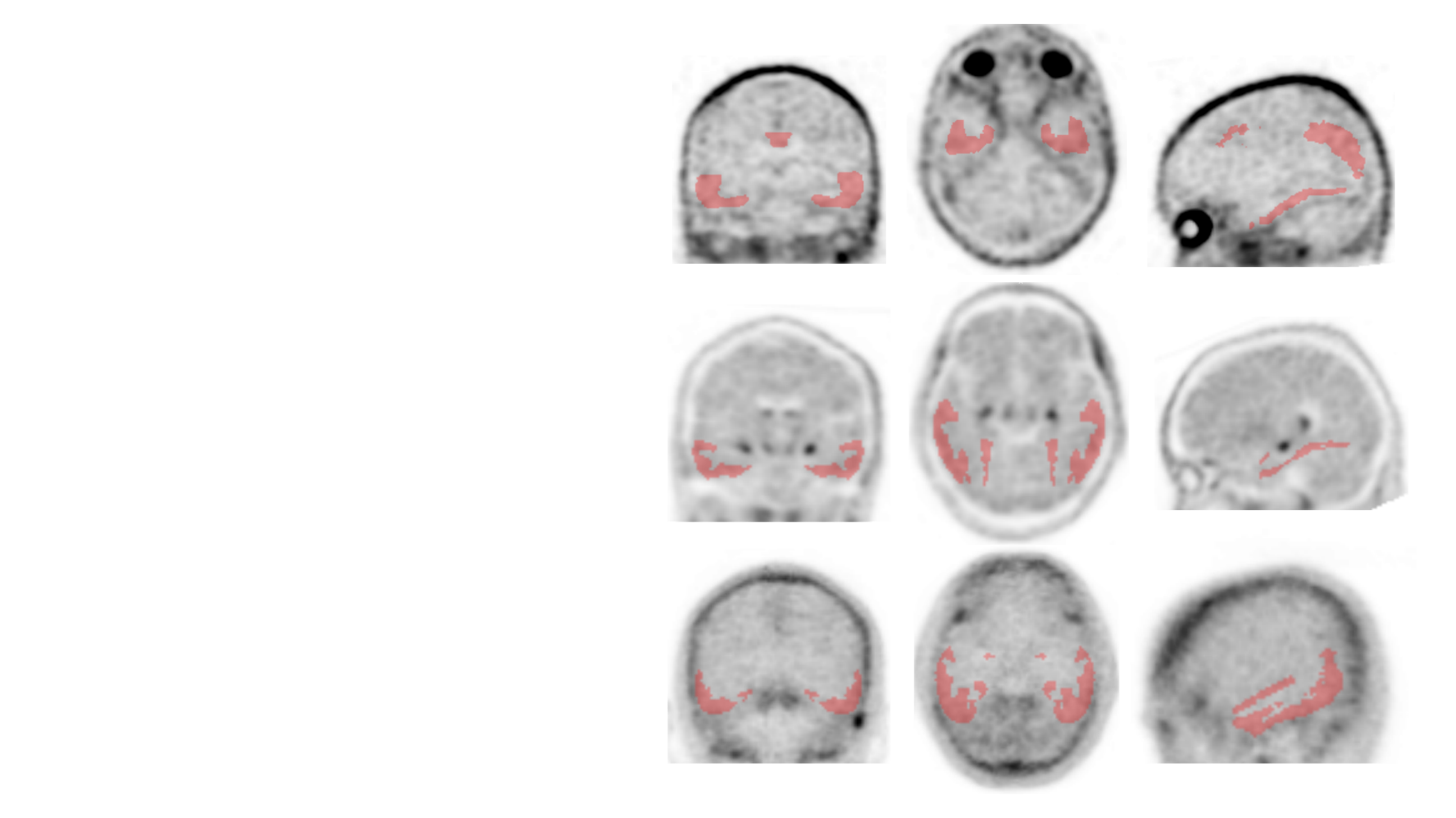

**Supplementary Figure 4:** boxplot of the SPM CTR_z_ between CU Aβ- and AD Aβ+ for the 6 different tau tracers. The blue dashed line corresponds to 2 CTRz.

Mesial Temporal

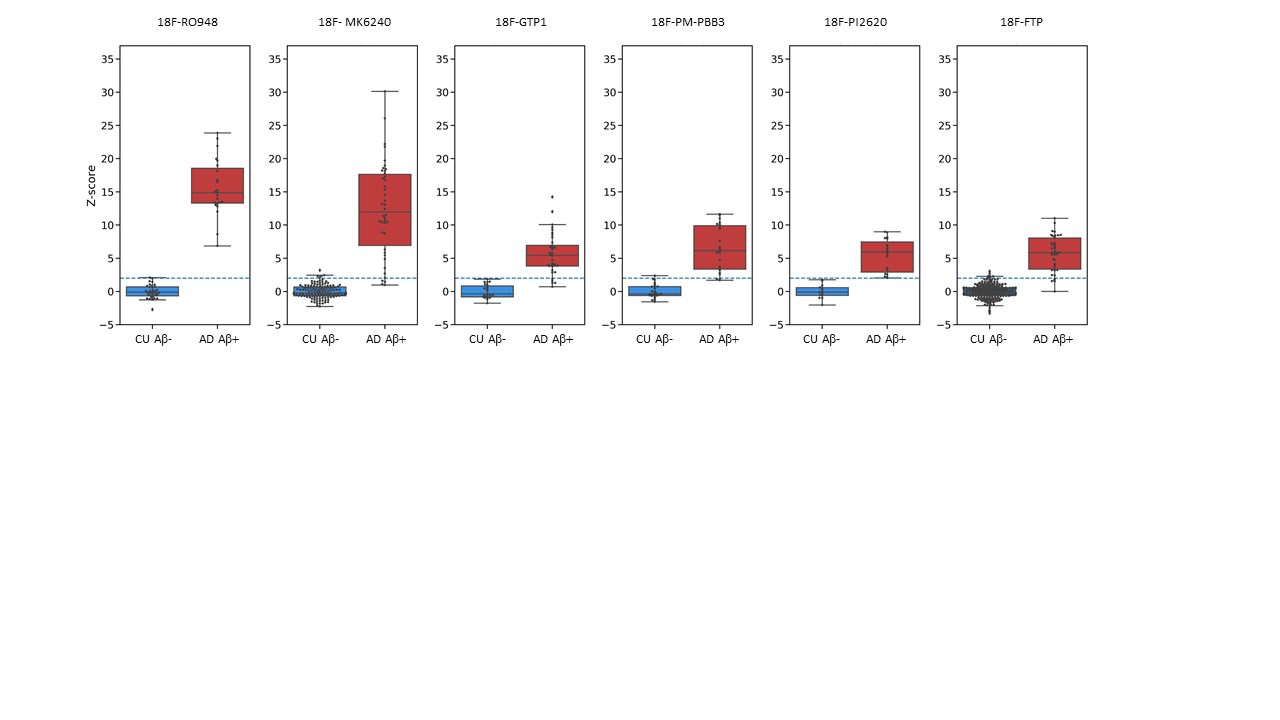

Temporo-Parietal

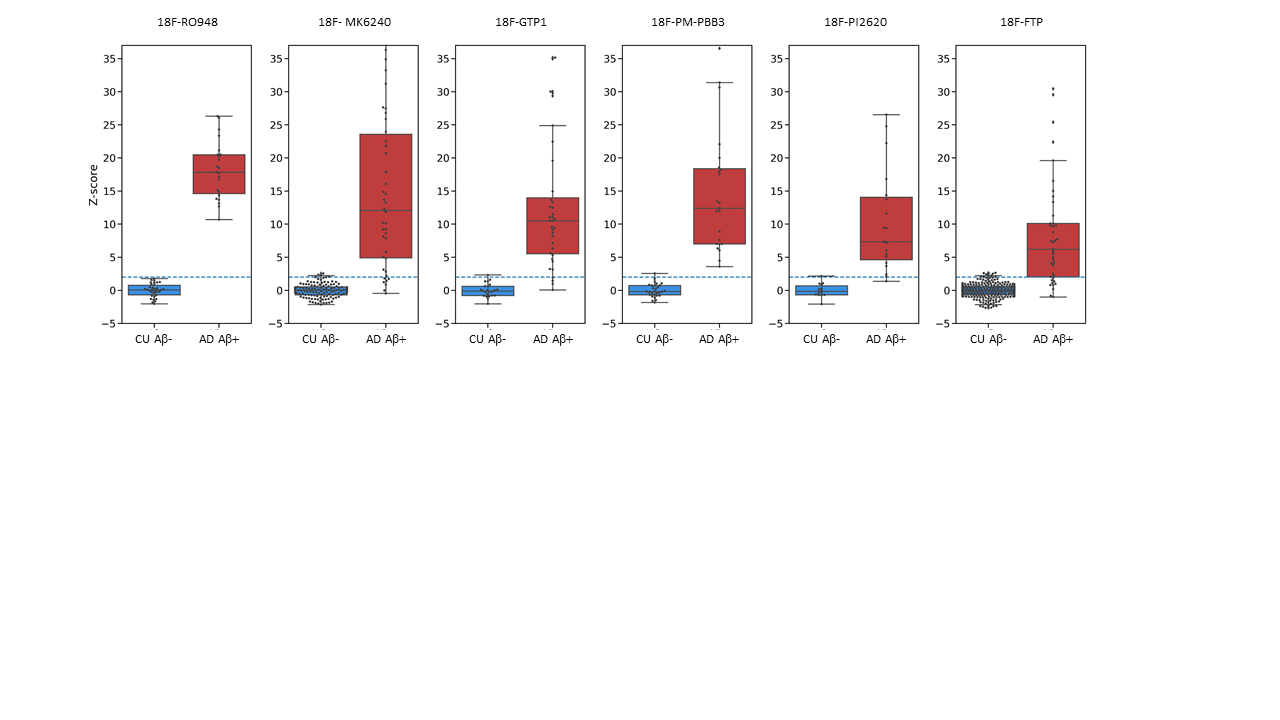

Frontal

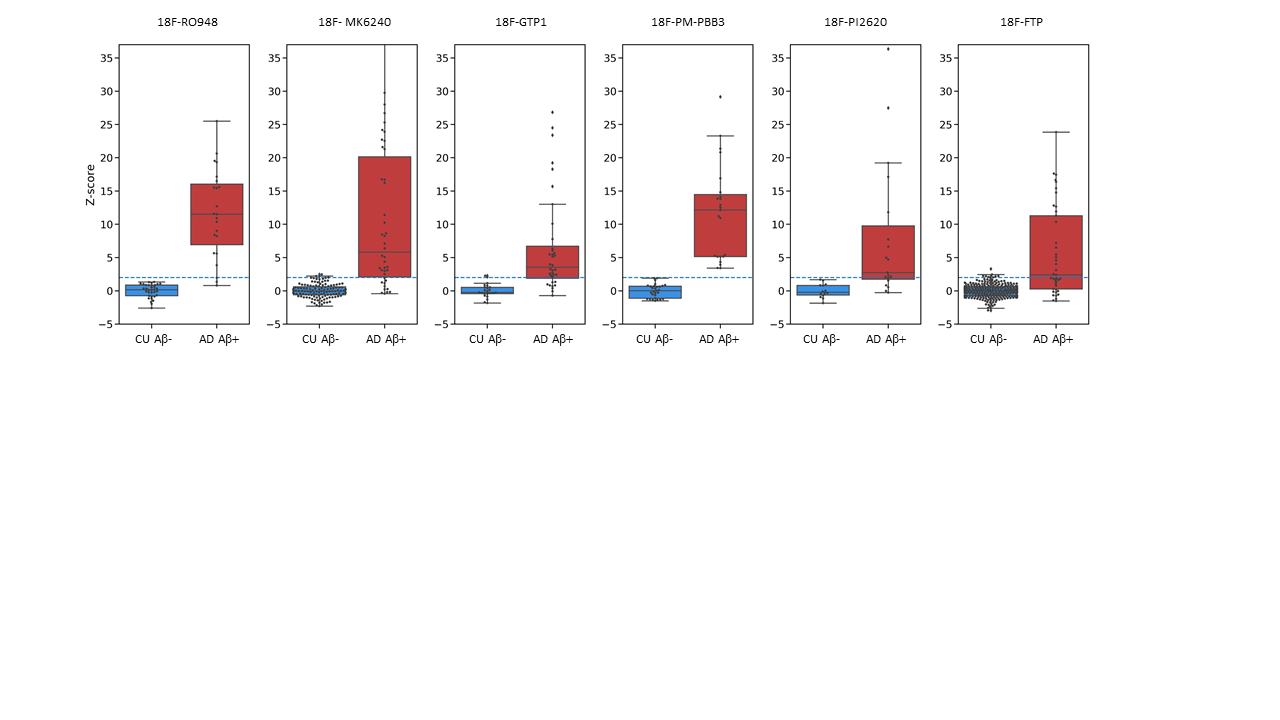

Global

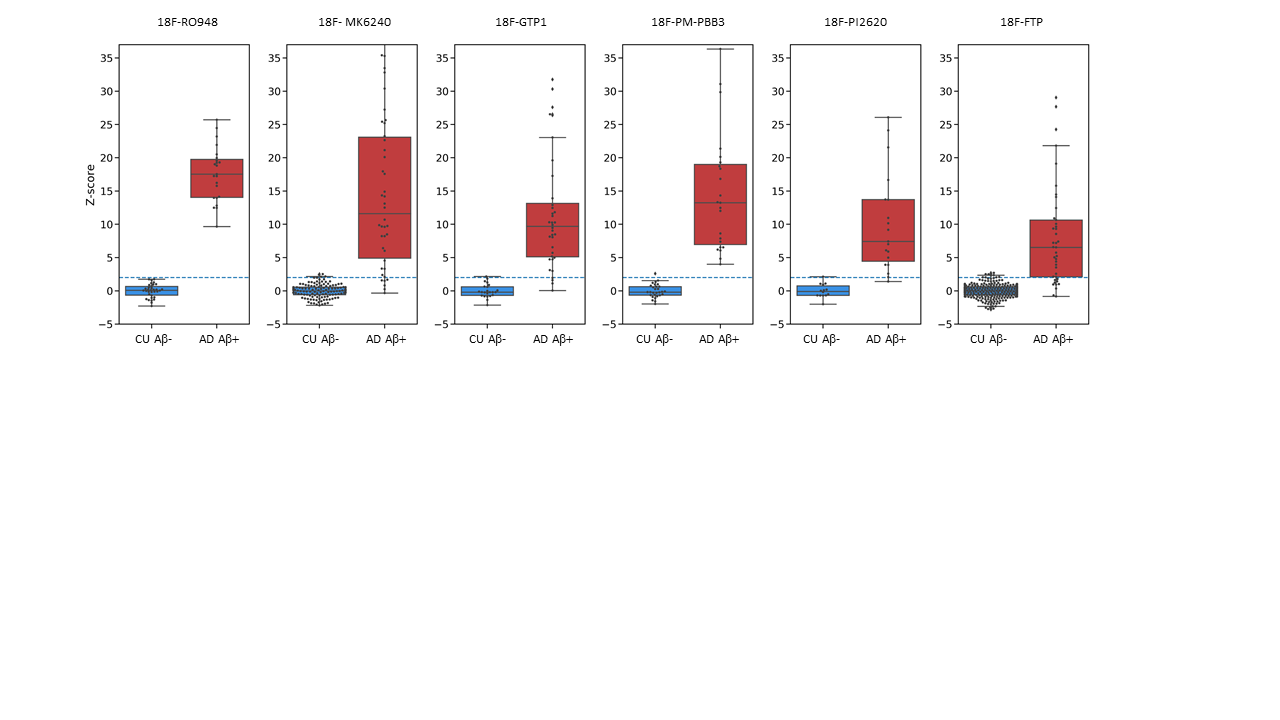

**Supplementary Figure 5:** ROC analysis of ROI CTR_z_ to discriminate AD A*β*+ from other sub-cohorts. Black dashed lines indicate the sensitivities and specificities at 2 CTR_z_

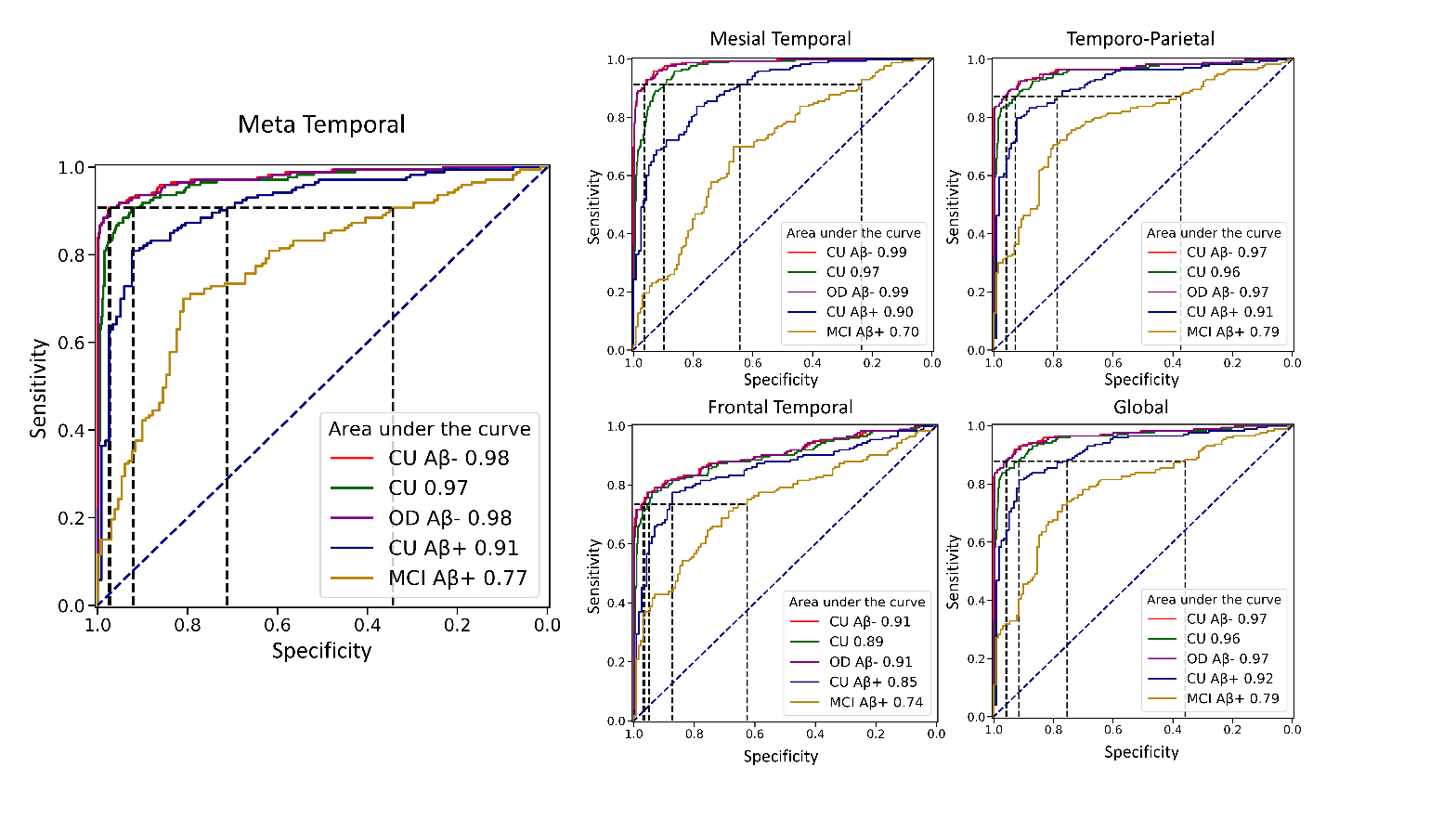

**Supplementary Figure 6:** boxplot of the CapAIBL CTR_z_ between CU Aβ- and AD Aβ+ for the 6 different tau tracers. The blue dashed line corresponds to 2 CTRz.

Mesial Temporal

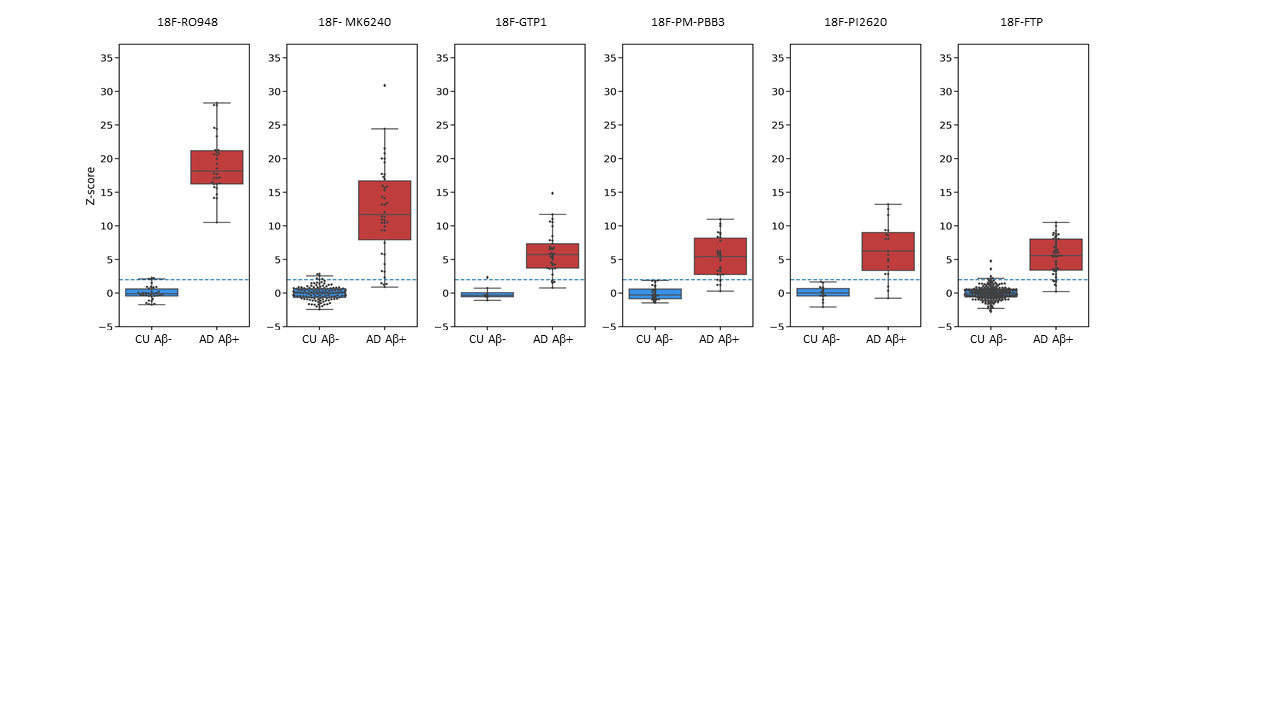

Meta Temporal

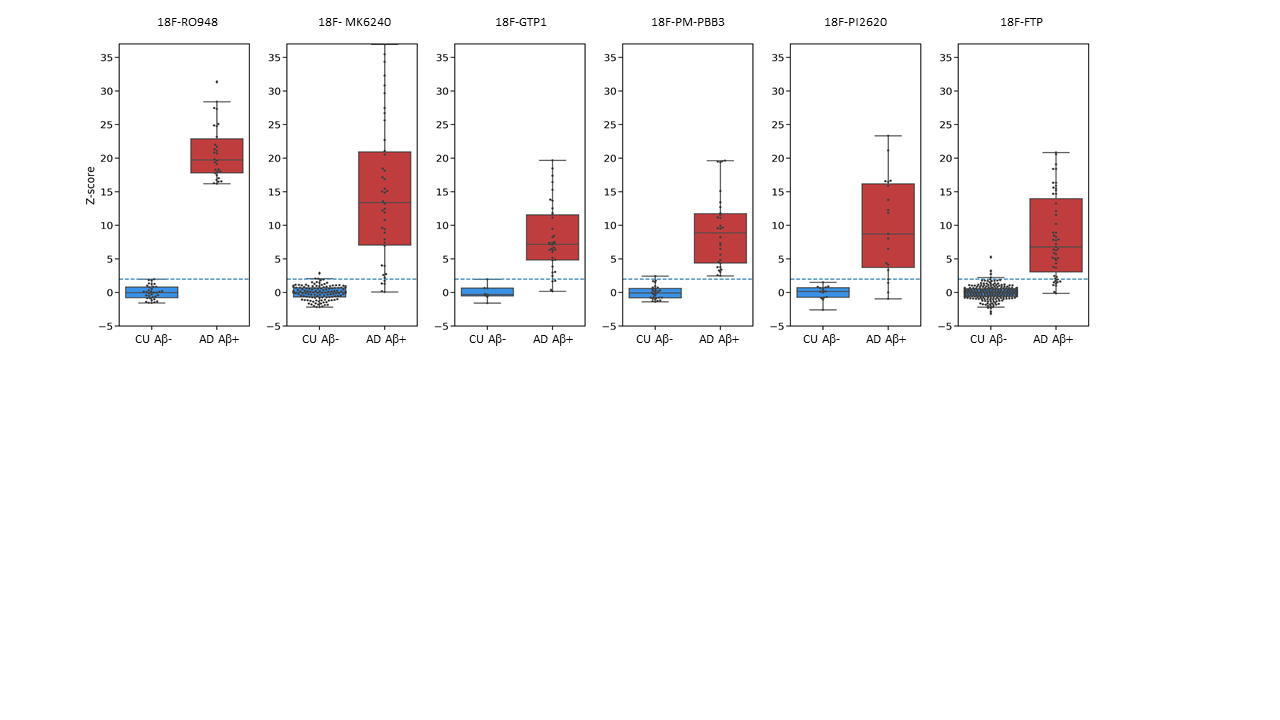

Temporo-Parietal

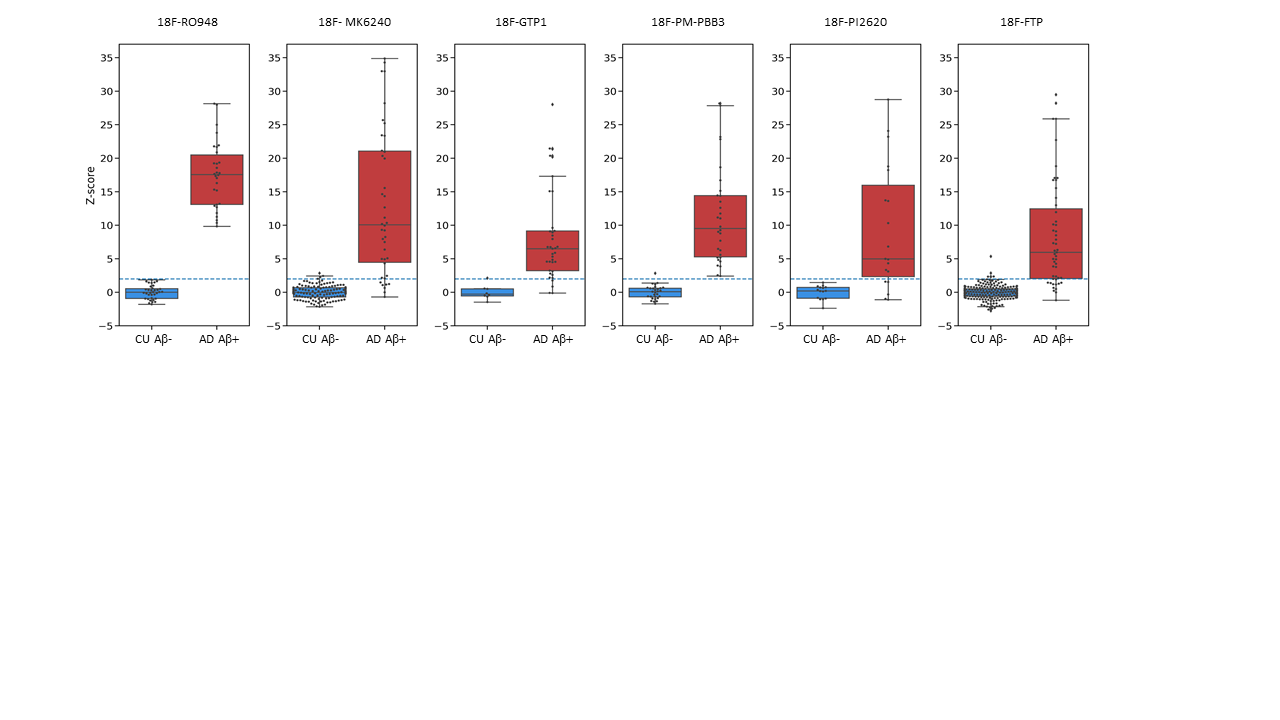

Frontal

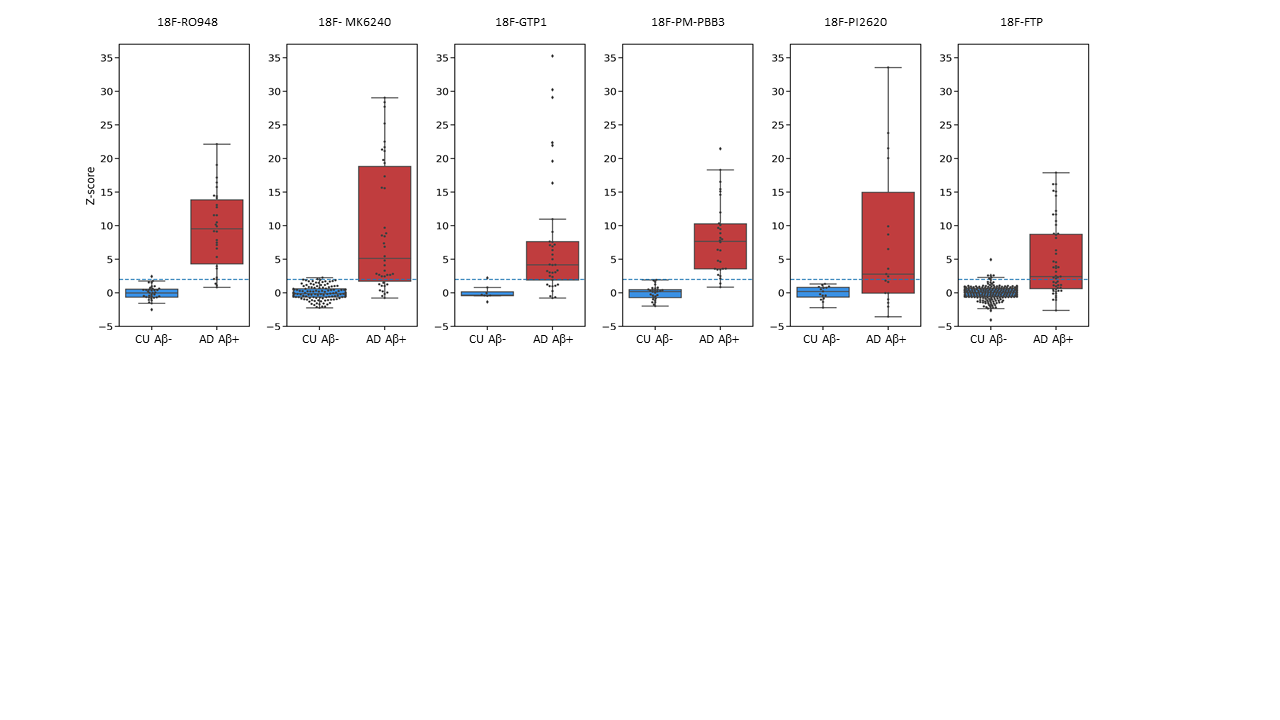

Global

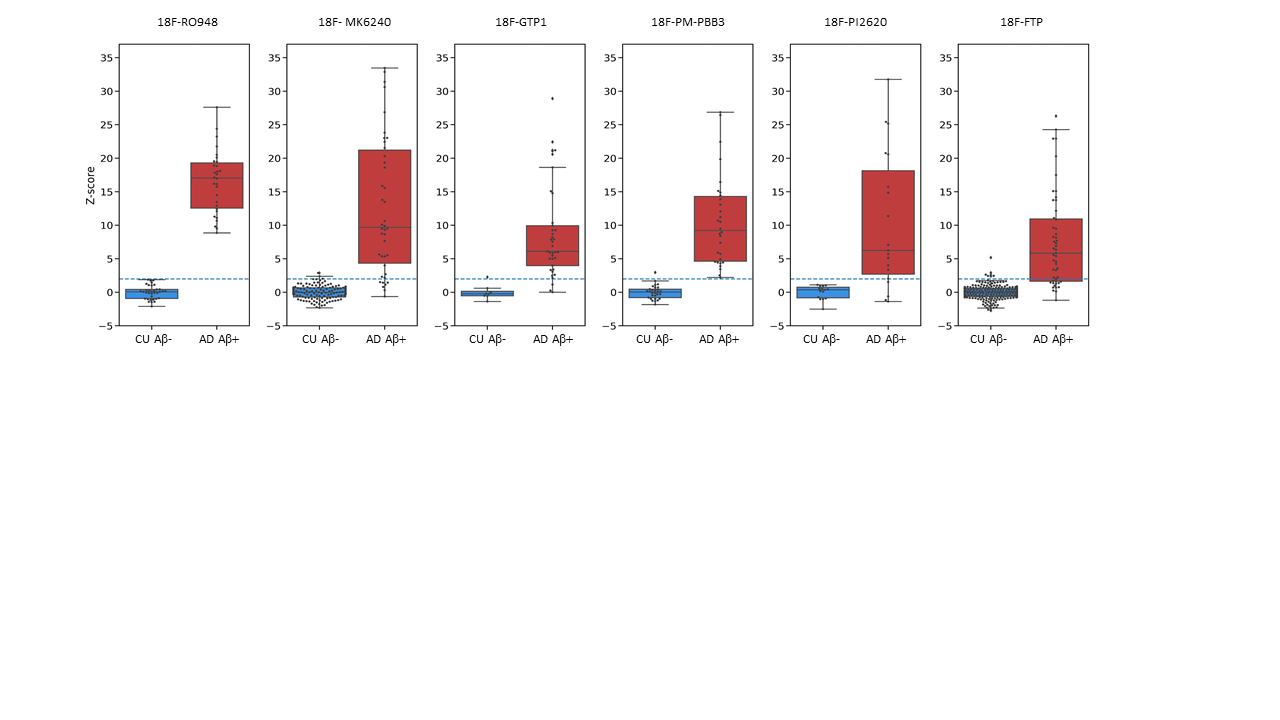

**Supplementary Figure 7:** ROC analysis of CapAIBL ROI CTR_z_ to discriminate AD Aβ+ from other sub-cohorts. Black dashed lines indicate the sensitivities and specificities at 2 CTR_z_.

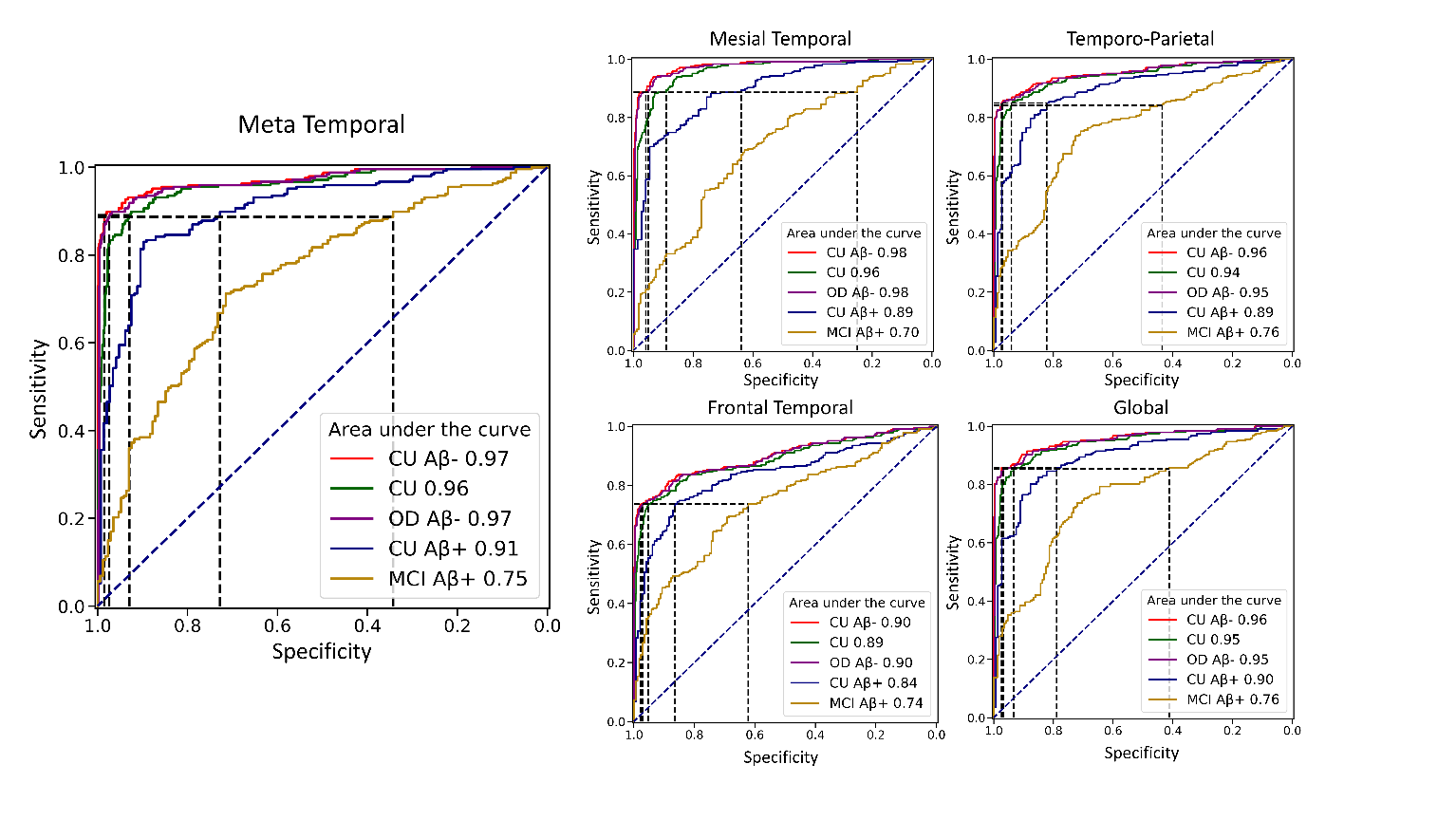

**Supplementary Figure 8:** Boxplots of the ROI CTR_z_ in the different ROIs.

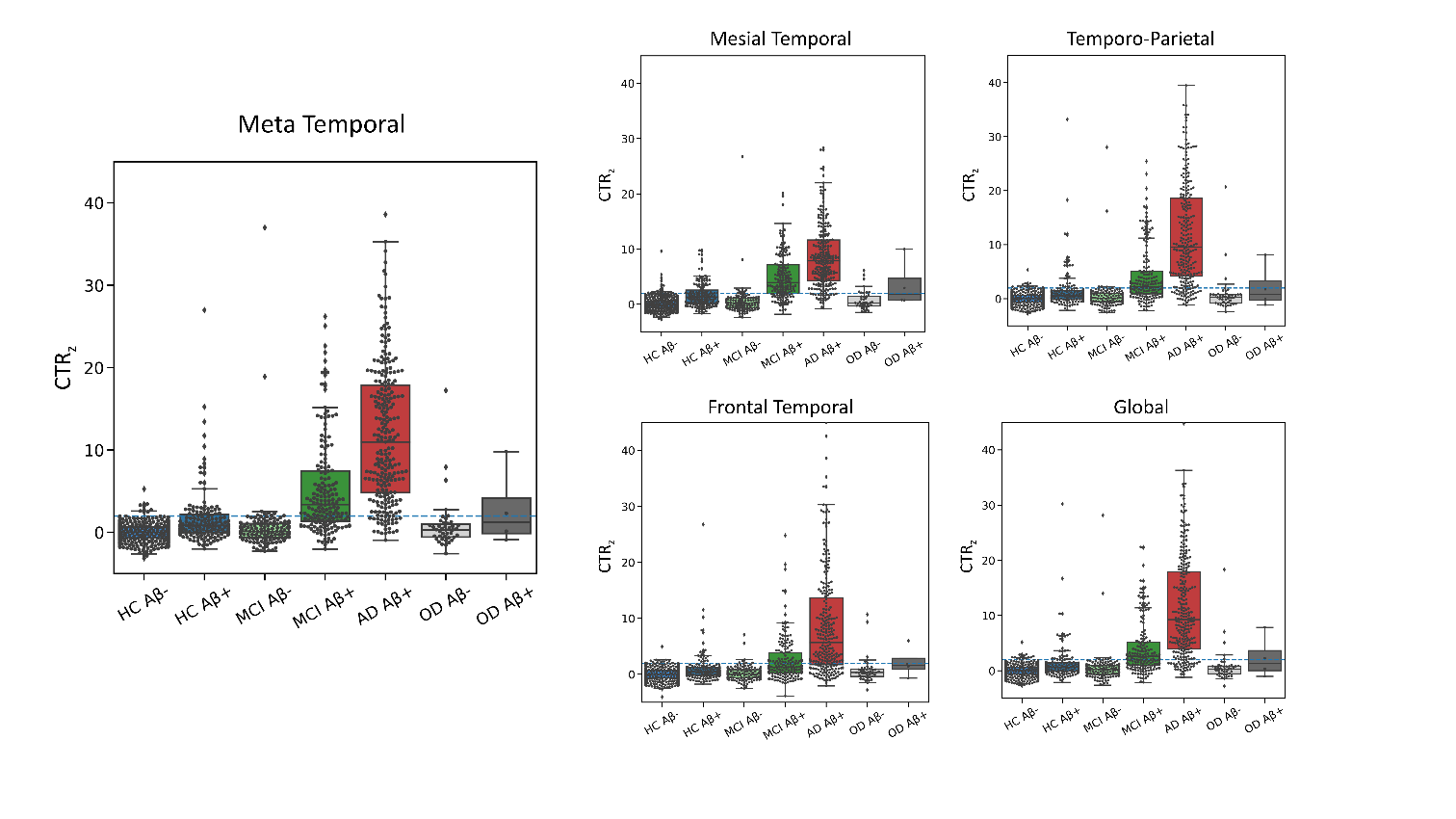

**Supplementary Figure 9:** SPM ROI CTR_z_ as a function of centiloid. A CTR_z_ higher than 2 in the cortex (in the frontal) is rare in individuals with a CL lower than 50CL (70CL). The blue dashed line corresponds to 2 CTRz.

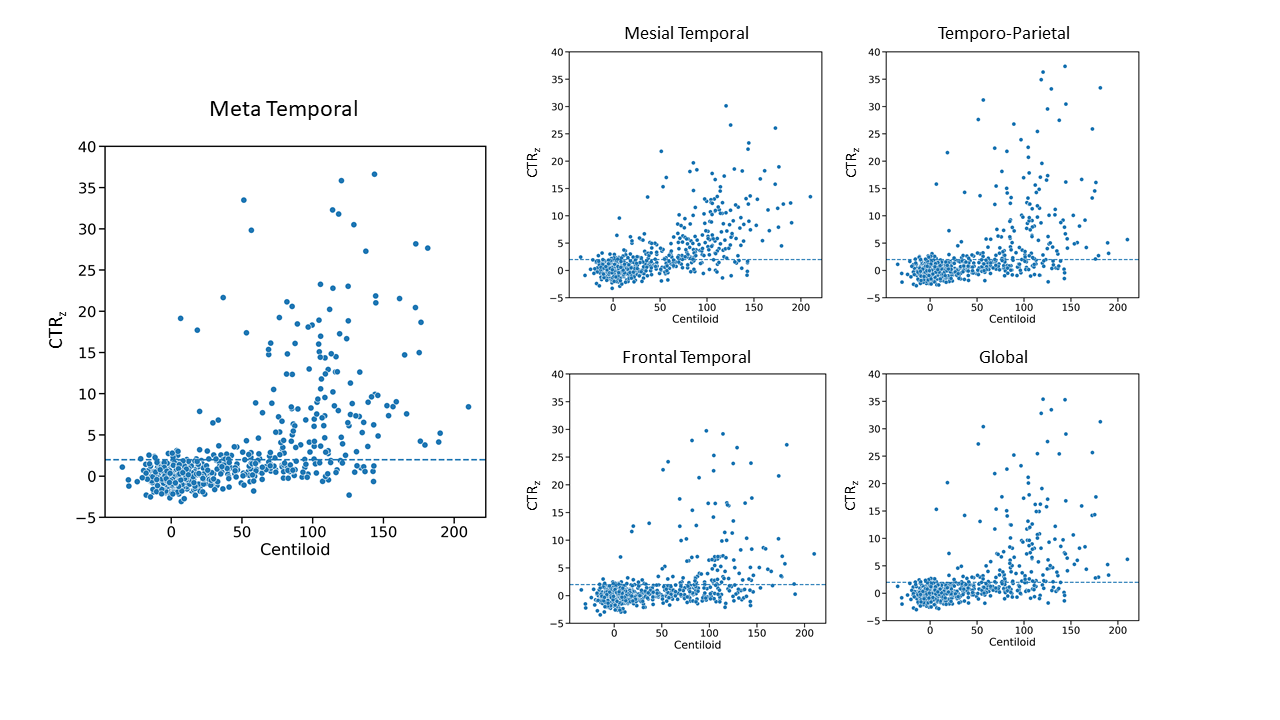

**Supplementary Figure 10:** CapAIBL ROI CTR_z_ as a function of centiloid. A CTR_z_ higher than 2 in the cortex (in the frontal) is rare in individuals with a CL lower than 50CL (70CL). The blue dashed line corresponds to 2 CTRz.

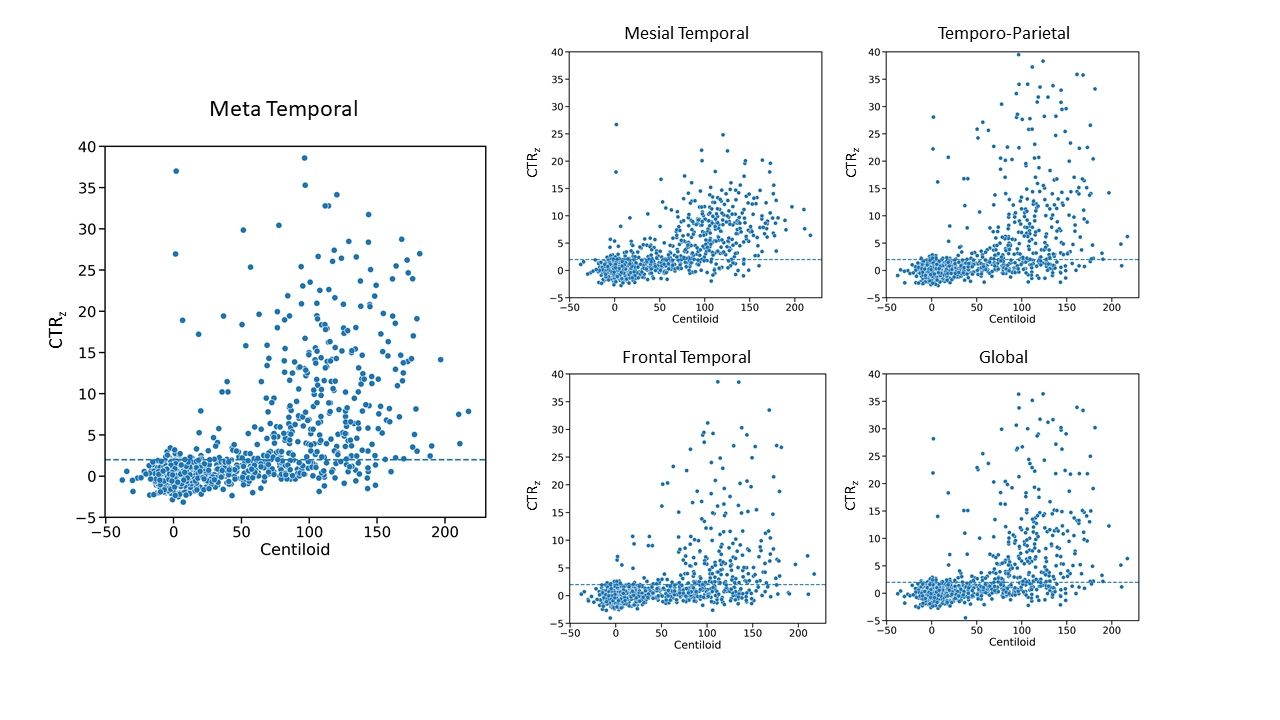

**Supplementary Figure 11:** Scatter plots of the CapAIBL CTR_z_ in the Meta Temporal and temporo-Parietal as a function of the CapAIBL CTR_z_ in the Mesial temporal. Points are coloured depending on their visual reads. The blue dashed lines correspond to 2 CTRz.

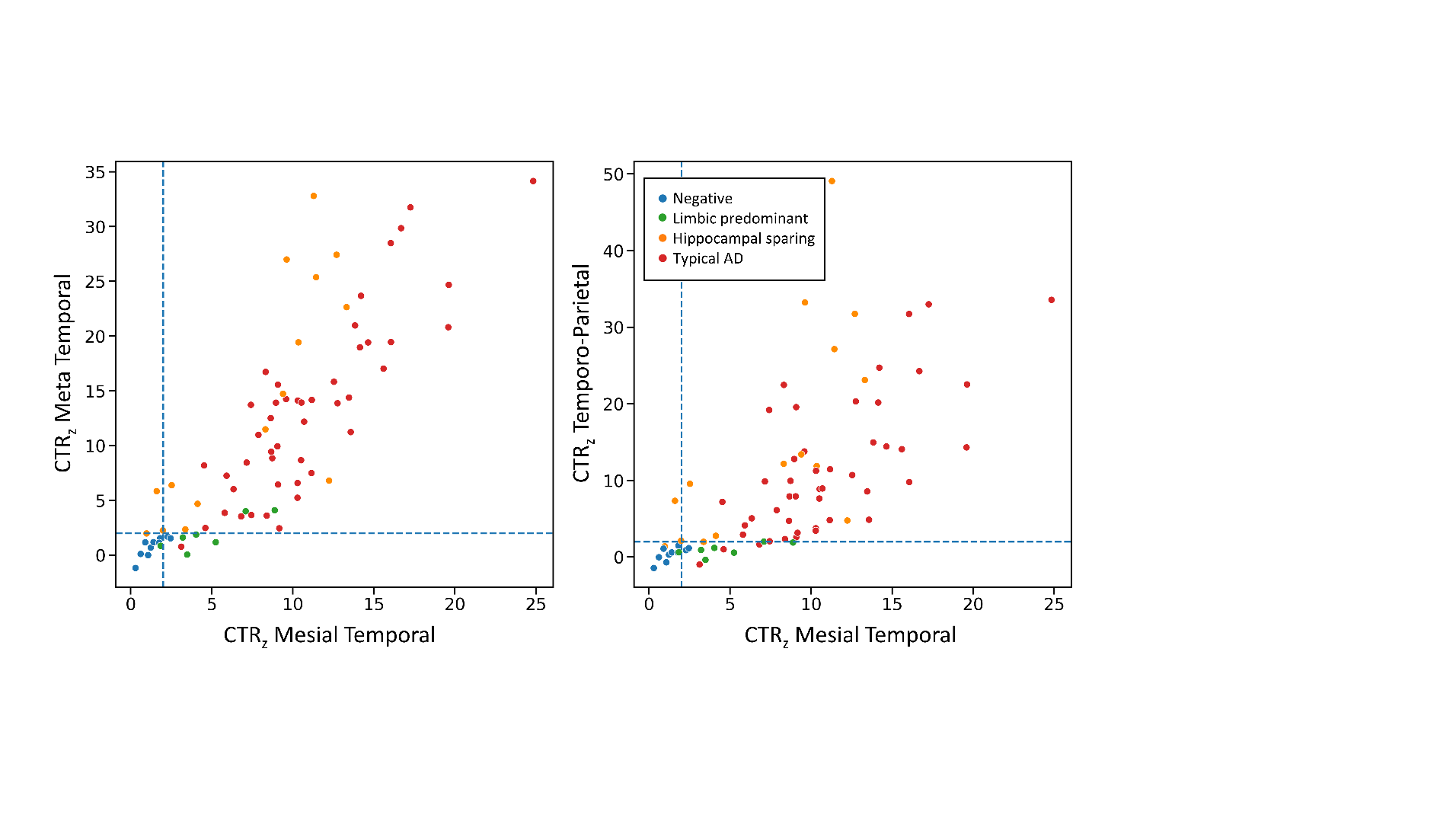

**Supplementary Figure 12:** Surface projections of 4 AD Aβ+ ^18^F-MK6240 scans from 4 visual tau topological patterns, tau negative, Limbic predominant, hippocampal sparing and typical. The scale is displayed in SUVR and CTR_z_.

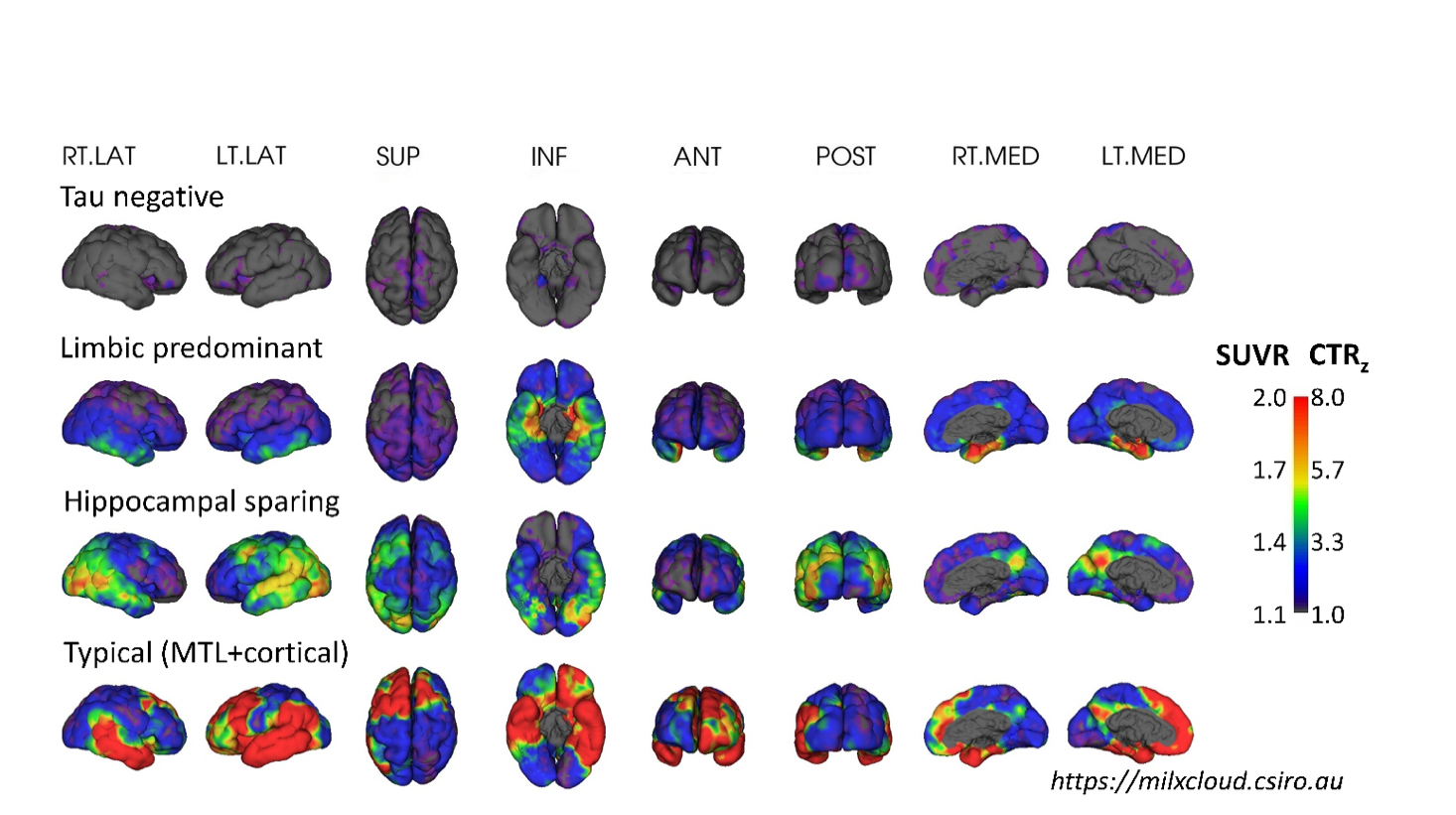

1. *The ADNI was launched in 2003 as a public-private partnership, led by Principal Investigator Michael W. Weiner, MD. The primary goal of ADNI has been to test whether serial magnetic resonance imaging (MRI), positron emission tomography (PET), other biological markers, and clinical and neuropsychological assessment can be combined to measure the progression of mild cognitive impairment (MCI) and early Alzheimer's disease (AD). Tau and amyloid acquisition information can be found on the ADNI page, see www.adni-info.org*.
